# Reproducible And Clinically Translatable Deep Neural Networks For Cervical Screening

**DOI:** 10.1101/2022.12.17.22282984

**Authors:** Syed Rakin Ahmed, Brian Befano, Andreanne Lemay, Didem Egemen, Ana Cecilia Rodriguez, Sandeep Angara, Kanan Desai, Jose Jeronimo, Sameer Antani, Nicole Campos, Federica Inturrisi, Rebecca Perkins, Aimee Kreimer, Nicolas Wentzensen, Rolando Herrero, Marta del Pino, Wim Quint, Silvia de Sanjose, Mark Schiffman, Jayashree Kalpathy-Cramer

**Affiliations:** Athinoula A. Martinos Center for Biomedical Imaging, Department of Radiology, Massachusetts General Hospital, Boston, MA 02129, USA; Harvard Graduate Program in Biophysics, Harvard Medical School, Harvard University, Cambridge, MA 02115, USA; Massachusetts Institute of Technology, Cambridge, MA 02139, USA; Geisel School of Medicine at Dartmouth, Dartmouth College, Hanover, NH 02139,USA; Information Management Services, Calverton, MD 20705, USA; University of Washington, Seattle, WA 98195, USA; NeuroPoly, Polytechnique Montreal, Montreal, QC H3T 1N8, Canada; Clinical Epidemiology Unit, Clinical Genetics Branch, Division of Cancer Epidemiology and Genetics, National Cancer Institute, National Institutes of Health, Bethesda, MD 20892; Computational Health Research Branch, National Library of Medicine, Lister Hill Center, Bethesda, MD 20894; Department of Health Policy and Management, Harvard T.H. Chan School of Public Health, Boston MA 02115; Dept of Obstetrics & Gynecology, Boston University Chobanian & Avedisian School of Medicine, Boston, MA 02118; Agencia Costarricense de Investigaciones Biomedicas (ACIB), Fundacion INCIENSA, San Jose, Costa Rica; Hospital Clinic, Barcelona, Spain; DDL Diagnostic Laboratory, Rijswijk, The Netherlands; ISGlobal, Barcelona, Spain

**Keywords:** human papillomavirus, cervical cancer screening, artificial intelligence, deep learning

## Abstract

Cervical cancer is a leading cause of cancer mortality, with approximately 90% of the 250,000 deaths per year occurring in low- and middle-income countries (LMIC). Secondary prevention with cervical screening involves detecting and treating precursor lesions; however, scaling screening efforts in LMIC has been hampered by infrastructure and cost constraints. Recent work has supported the development of an artificial intelligence (AI) pipeline on digital images of the cervix to achieve an accurate and reliable diagnosis of treatable precancerous lesions. In particular, WHO guidelines emphasize visual triage of women testing positive for human papillomavirus (HPV) as the primary screen, and AI could assist in this triage task. Published AI reports have exhibited overfitting, lack of portability, and unrealistic, near-perfect performance estimates. To surmount recognized issues, we implemented a comprehensive deep-learning model selection and optimization study on a large, collated, multi-institutional dataset of 9,462 women (17,013 images). We evaluated relative portability, repeatability, and classification performance. The top performing model, when combined with HPV type, achieved an area under the Receiver Operating Characteristics (ROC) curve (AUC) of 0.89 within our study population of interest, and a limited total extreme misclassification rate of 3.4%, on held-aside test sets. Our work is among the first efforts at designing a robust, repeatable, accurate and clinically translatable deep-learning model for cervical screening.

---

The flood of artificial intelligence (AI) and deep learning (DL) approaches in recent years (1,2) has permeated medicine and medical imaging, where it has had a transformative impact: some AI based algorithms are now able to interpret imaging at the level of experts (3,4). This can be attributed to three key factors: 1. a pressing and seemingly consistent clinical need; 2. the advancements in and convergence of computational resources, innovations, and collaborations; and 3. the generation of larger and more comprehensive repositories of patient image data for model development (5). The nature of clinical tasks performed by AI models has shifted from simple detection or classification to more nuanced versions with direct relevance for risk stratification of patients and precision medicine (6).

The advancements made by AI in image classification tasks over the past several years have also reached the cervical imaging domain, for instance, as an assistive technology for cervical screening (7). Globally, cervical cancer is a leading cause of cancer morbidity and mortality, with approximately 90% of the 250,000 deaths per year occurring in low- and middle-income countries (LMIC) (8,9). Persistent infections with high-risk human papillomavirus (HPV) types are the causal risk factor for subsequent carcinogenesis (10,11). Accordingly, primary prevention via prophylactic HPV vaccination (12), and secondary prevention via HPV-based screening for precursor lesions (“precancer”) are the recommended preventive methods (13,14). Crucially, screening is the key secondary prevention strategy, with the long process of carcinogenic transformation from HPV infection to invasive cancer providing an opportunity for detecting the disease at a stage when treatment is preventive or, at least, curative (13).

However, implementation of an effective cervical screening program in LMIC, in line with WHO’s elimination targets (15), is hindered by barriers to healthcare delivery. Cytology and other current tests are costly and have substantial infrastructure requirements due to the need for laboratory infrastructure, transport of samples, multiple visits for screening and treatment, and (in the case of cytology) highly trained cytopathologists and colposcopists for management of abnormal results (16). As a less resource-intensive alternative, some have established screening of the cervix by visual inspection after application of acetic acid (VIA) to identify precancerous or cancerous abnormalities via community-based programs, followed by treatment of abnormal lesions using thermal ablation or cryotherapy and/or large loop excision of the transformation zone (LLETZ) (17,18). The major limitation of VIA, however, is its inherently subjective and unreliable nature, resulting in high variability in the ability of clinicians to differentiate precancer from more common minor abnormalities, which leads to both undertreatment and overtreatment (19,20).

Given the severe burden of cervical cancer and the lack of widely disseminated screening approaches in LMIC, a critical need exists for methods that can more consistently, inexpensively, and accurately evaluate cervical lesions and subsequently enable informed local choice of the appropriate treatment protocols.

There has been a relative paucity of prior work utilizing AI and DL for cervical screening based on cervical images. Crucially, the existing work also largely suffers from overfitting of the model on the training data. This leads to apparent initial promise, with either poor performance on or absence of held-aside test sets for evaluating true model performance. When deployed in different settings, these models fail to return consistent scores and accurately detect precancers (21–24). This poses significant concerns when considering downstream deployment in various LMIC, where model predictions directly inform the course of treatment, and where screening opportunities are limited.

In this work, we address the aforementioned concerns through three contributions, which are generalizable to clinical domains outside of cervical imaging:

1. <u>Improved reliability of model predictions</u> We employ a comprehensive, multi-level model design approach with a primary aim of improving model reliability. Model reliability or repeatability, is defined as the ability of a model to generate near-identical predictions for the same woman under identical conditions, ensuring that the model produces precise, reliable outputs in the clinical setting. Specifically, we consider multiple combinations of model architectures, loss functions, balancing strategies, and dropout. Our final model selection for the classifier, termed automated visual evaluation (AVE), is based on a criterion that first prioritizes model reliability, followed by class discrimination or classification performance, and finally reduction of grave errors.
2. <u>Improved clinical translatability: multi-level ground truth</u> The large majority of current medical image classification and radiogenomic pipelines that utilize AI and DL, across clinical domains, use binary ground truths. Our clinical intuition from working with binary models as well as prior empirical work have informed us that these models frequently fail to capture the inherent uncertainty with ambiguous samples (21–24). These uncertain samples are of two intersecting kinds: samples that are uncertain to the clinician (“rater uncertainty”) and samples that are uncertain to the model i.e., where the model reports low confidence scores (“model uncertainty”); both instances can lead to incorrect classification and subsequent misinformed downstream actions for these patients. Crucially, real-world clinical oncology samples, across domains such as cervical, prostate and breast, and across hospitals/institutions, include many uncertain cases (25–27). To address both levels of ambiguity, we employ several multi-level, ordinal ground truth delineation schemes in our model selection.
3. <u>Improved downstream clinical-decision making: combination of HPV risk stratification with model predictions</u> A number of different cancers have identified “sufficient” causes. Examples across this spectrum range from the presence of BRAF V600E mutation for the papillary subtype for craniopharyngioma (28), to the presence of BRCA1 or BRCA2 mutations for breast cancer (29–31). Cervical cancer is unique among common neoplasms in that HPV is virtually necessary and is present in >95% of cases. Different HPV types predict higher or lower absolute risk, e.g., HPV 16 is the highest risk type, followed by HPV 18, while other types pose weaker or no risk (32–34). In our work, we combined HPV typing and its strong risk stratification with our visual model predictions, to create a risk score that can be adapted to local clinical preferences for “risk-action” thresholds. This is generalizable across clinical domains where additional clinical variables and risk associations significantly determine patient outcomes.

## RESULTS

In this work, we conducted a comprehensive, multi-stage model selection and optimization approach (Fig. 1, Fig. 2), utilizing a large, collated multi-institution, multi-device, and multi-population dataset of 9,462 women (17,013 images) (Table 1), in order to generate a diagnostic classifier optimized for 1. repeatability; 2. classification performance; and 3. HPV-group combined risk stratification (Fig. 2) (see METHODS).

**FIGURE 1:**
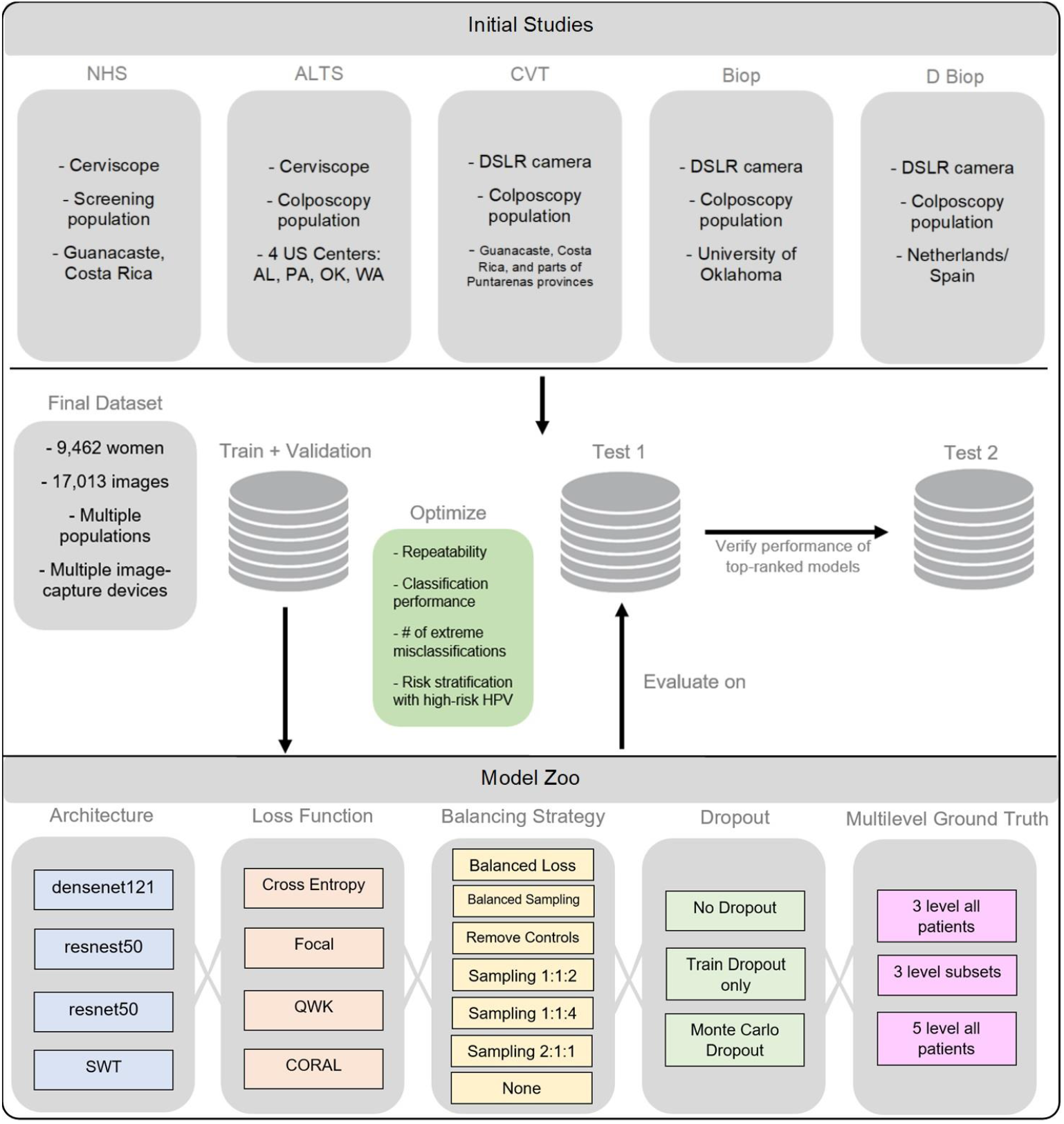
Model selection and optimization overview. The top panel highlights the five different studies (NHS, ALTS, CVT, Biop and D Biop; see Table 1, Supp. Table 1, and Supp. Methods for detailed description and breakdown of the studies by ground truth) used to generate the final dataset on the middle panel, which is subsequently used to generate a train and validation set, as well as two separate test sets. The intersections of model selection choices on the bottom panel are used to generate a compendium of models trained using the corresponding train and validation sets and evaluated on Test Set 1, optimizing for repeatability, classification performance, reduced extreme misclassifications and combined risk-stratification with high-risk human papillomavirus (HPV) types. Test Set 2 is utilized to verify the performance of top candidates that emerge from evaluation on Test Set 1. SWT: Swin Transformer; QWK: quadratic weighted kappa; CORAL: CORAL (consistent rank logits) loss, as described in the METHODS section.

**FIGURE 2:**
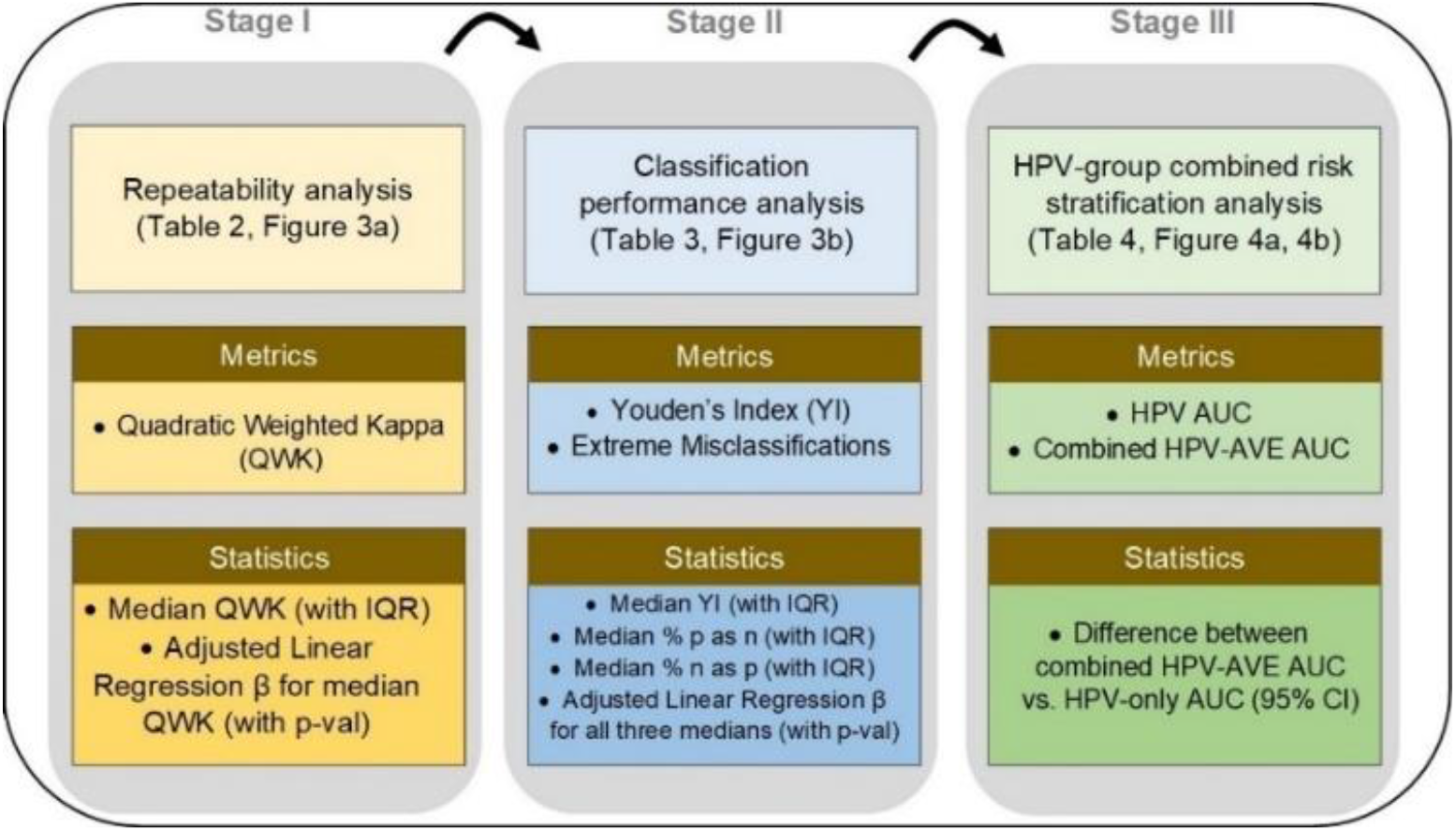
Model selection approach and statistical analysis utilized in our automated visual evaluation (AVE) classifier. IQR: interquartile range; AUC: area under the receiver operating characteristics (ROC) curve; CI: confidence interval.

**TABLE 1:** Baseline characteristics of women in each of the ground truth categories, highlighting proportions by histology, cytology, human papillomavirus (HPV) type, study, as well as age and # images/woman. The detailed study descriptions and ground truth assignment by study can be found in Supp. Table 1 and in the Supp. Methods section. CIN: cervical intraepithelial neoplasia; AIS: adenocarcinoma in situ; ASC-H: atypical squamous cells, cannot rule out high grade squamous intraepithelial lesion; HSIL: high-grade squamous intraepithelial lesion; LSIL: low-grade squamous intraepithelial lesion; ASCUS: atypical squamous cells of undetermined significance; SD: standard deviation; IQR: interquartile range.

| Table 1: Baseline characteristics of women in each of the ground truth categories |  |  |  |  |  |  |  |  |  |  |
| --- | --- | --- | --- | --- | --- | --- | --- | --- | --- | --- |
| Characteristics | Ground truth categories |  |  |  |  |  |  |  |  |  |
|  | no. (%) |  |  |  |  |  |  |  |  |  |
|  | Normal<br>(N=6092) |  | Gray Low<br>(N=867) |  | Gray Middle<br>(N=918) |  | Gray High<br>(N=529) |  | Precancer+<br>(N=1056) |  |
| **Histology** |  |  |  |  |  |  |  |  |  |  |
| Cancer | 0 | (0.0%) | 0 | (0.0%) | 0 | (0.0%) | 0 | (0.0%) | 23 | (2.2%) |
| CIN3/AIS | 0 | (0.0%) | 0 | (0.0%) | 0 | (0.0%) | 0 | (0.0%) | 571 | (54.1%) |
| CIN2 | 0 | (0.0%) | 0 | (0.0%) | 1 | (0.1%) | 66 | (12.5%) | 456 | (43.2%) |
| <CIN2 | 873 | (14.3%) | 467 | (53.9%) | 580 | (63.2%) | 280 | (52.9%) | 6 | (0.6%) |
| No histology | 5219 | (85.7%) | 400 | (46.1%) | 337 | (36.7%) | 183 | (34.6%) | 0 | (0.0%) |
| **Cytology** |  |  |  |  |  |  |  |  |  |  |
| ASC-H/HSIL | 0 | (0.0%) | 164 | (18.9%) | 110 | (12.0%) | 481 | (90.9%) | 647 | (61.3%) |
| LSIL | 0 | (0.0%) | 220 | (25.4%) | 586 | (63.8%) | 15 | (2.8%) | 209 | (19.8%) |
| ASCUS | 4288 | (70.4%) | 95 | (11.0%) | 222 | (24.2%) | 19 | (3.6%) | 112 | (10.6%) |
| Normal | 1801 | (29.6%) | 386 | (44.5%) | 0 | (0.0%) | 11 | (2.1%) | 67 | (6.3%) |
| Other/missing | 3 | (0.0%) | 2 | (0.2%) | 0 | (0.0%) | 3 | (0.6%) | 21 | (2.0%) |
| **HPV type** |  |  |  |  |  |  |  |  |  |  |
| 16 | 0 | (0.0%) | 95 | (11.0%) | 172 | (18.7%) | 174 | (32.9%) | 507 | (48.0%) |
| 18, 45 | 0 | (0.0%) | 66 | (7.6%) | 141 | (15.4%) | 54 | (10.2%) | 123 | (11.6%) |
| 31,33,35,52,58 | 0 | (0.0%) | 187 | (21.6%) | 346 | (37.7%) | 174 | (32.9%) | 312 | (29.5%) |
| 39,51,56,59,68 | 0 | (0.0%) | 130 | (15.0%) | 250 | (27.2%) | 59 | (11.2%) | 78 | (7.4%) |
| Negative | 6087 | (99.9%) | 382 | (44.1%) | 6 | (0.7%) | 68 | (12.9%) | 26 | (2.5%) |
| Missing | 5 | (0.1%) | 7 | (0.8%) | 3 | (0.3%) | 0 | (0.0%) | 10 | (0.9%) |
| **Study** |  |  |  |  |  |  |  |  |  |  |
| NHS | 4518 | (74.2%) | 114 | (13.1%) | 127 | (13.8%) | 34 | (6.4%) | 173 | (16.4%) |
| ALTS | 943 | (15.5%) | 231 | (26.6%) | 314 | (34.2%) | 171 | (32.3%) | 363 | (34.4%) |
| CVT | 424 | (7.0%) | 297 | (34.3%) | 208 | (22.7%) | 49 | (9.3%) | 195 | (18.5%) |
| Biop | 66 | (1.1%) | 51 | (5.9%) | 63 | (6.9%) | 32 | (6.0%) | 132 | (12.5%) |
| D Biop | 141 | (2.3%) | 174 | (20.1%) | 206 | (22.4%) | 243 | (45.9%) | 193 | (18.3%) |
| **Age (30-49)** |  |  |  |  |  |  |  |  |  |  |
| Mean (SD) | 34.5 | (6.8) | 30.7 | (5.8) | 30.1 | (5.0) | 30.3 | (5.4) | 30.6 | (5.6) |
| Median (IQR) | 33 | (29-40) | 29 | (26-33) | 29 | (26-32) | 29 | (26-32) | 29 | (26-33) |
| **# images/woman** |  |  |  |  |  |  |  |  |  |  |
| Mean (SD) | 1.9 | (0.3) | 1.4 | (0.6) | 1.6 | (0.6) | 1.6 | (0.6) | 1.7 | (0.6) |
| Median (IQR) | 2 | (2-2) | 1 | (1-2) | 2 | (1-2) | 2 | (1-2) | 2 | (1-2) |

### REPEATABILITY ANALYSIS

Table 2 highlights the summary of the repeatability analysis (Stage I), reporting the mean, median and adjusted linear regression β values for QWK. We evaluated the metrics overall and within each design choice category, dropping the worst performing design choices both overall and within each category. Overall, this resulted in 19.0% of our design choices being dropped from further consideration (Table 2, shaded in salmon; Fig. 3a, muted bars). Within each design choice category, this amounted to dropping the design choices that had adjusted linear regression β values >0.06 below reference. Specifically, the design choices that were dropped in Stage 1 include the resnest50 architecture, focal and CORAL loss functions, and models trained without dropout. Here, we adopted a conservative approach, choosing to keep design choices that resulted in median QWK and corresponding adjusted β values that are relatively close and not clearly distinguishable from each other and only dropped the clearly worst performing choices; for instance, we decided to keep both the “3 level subsets” (β = - 0.026) and the “5 level all patients” (β = -0.025) design choices within the “Multilevel Ground Truth” design category, and pass them through to Stage 3.

**TABLE 2:** Repeatability analysis highlighting quadratic weighted kappa (QWK) summary statistics – mean, median with interquartile range (IQR) and adjusted linear regression (LR) β values – for design choices within each design choice category for our automated visual evaluation (AVE) classifier. Rows shaded in salmon indicate design choices filtered out at this stage due to poor repeatability. SWT: Swin Transformer; CORAL: CORAL (consistent rank logits) loss, as described in the METHODS section; ref: reference category.

| Table 2: Repeatability analysis |  |  |  |  |  |  |
| --- | --- | --- | --- | --- | --- | --- |
| Design Choice Category | Design Choices | QWK summary |  |  |  |  |
| | | Mean (SD) | | Median (IQR) | | Adjusted LR $\beta$ |
| Architecture | densenet121 | 0.743 | (0.062) | 0.748 | (0.719 - 0.786) | -0.016 |
|  | resnest50 | 0.675 | (0.069) | 0.649 | (0.630 - 0.743) | -0.083** |
|  | resnet50 | 0.752 | (0.048) | 0.760 | (0.736 - 0.776) | -0.018 |
|  | SWT | 0.743 | (0.079) | 0.748 | (0.671 - 0.815) | ref |
| Loss Function | Cross Entropy | 0.725 | (0.069) | 0.738 | (0.671 - 0.771) | -0.039** |
|  | Focal | 0.717 | (0.070) | 0.730 | (0.654 - 0.773) | -0.078** |
|  | QWK | 0.779 | (0.042) | 0.782 | (0.752 - 0.809) | ref |
|  | CORAL | 0.678 | (0.056) | 0.649 | (0.636 - 0.729) | -0.069** |
| Balancing strategy | Balanced loss | 0.703 | (0.107) | 0.751 | (0.647 - 0.769) | -0.053** |
|  | Balanced sampling | 0.729 | (0.057) | 0.735 | (0.675 - 0.781) | -0.046** |
|  | Remove controls | 0.775 | (0.054) | 0.777 | (0.744 - 0.809) | ref |
|  | Sampling 1:1:2 | 0.744 | (0.055) | 0.758 | (0.728 - 0.783) | -0.042** |
|  | Sampling 1:1:4 | 0.776 | (0.033) | 0.772 | (0.752 - 0.798) | -0.026 |
|  | Sampling 2:1:1 | 0.764 | (0.017) | 0.762 | (0.750 - 0.778) | -0.045 |
|  | None | 0.706 | (0.069) | 0.721 | (0.638 - 0.749) | -0.019 |
| Dropout | No Dropout | 0.663 | (0.072) | 0.649 | (0.620 - 0.723) | -0.088** |
|  | Train Dropout only | 0.725 | (0.058) | 0.738 | (0.681 - 0.759) | -0.035** |
|  | Monte Carlo Dropout | 0.760 | (0.059) | 0.772 | (0.733 - 0.802) | ref |
| Multilevel Ground Truth | 3 level all patients | 0.740 | (0.068) | 0.752 | (0.719 - 0.780) | ref |
|  | 3 level subsets | 0.707 | (0.070) | 0.709 | (0.637 - 0.778) | -0.026** |
|  | 5 level all patients | 0.705 | (0.064) | 0.721 | (0.650 - 0.748) | -0.025 |

**FIGURE 3:**
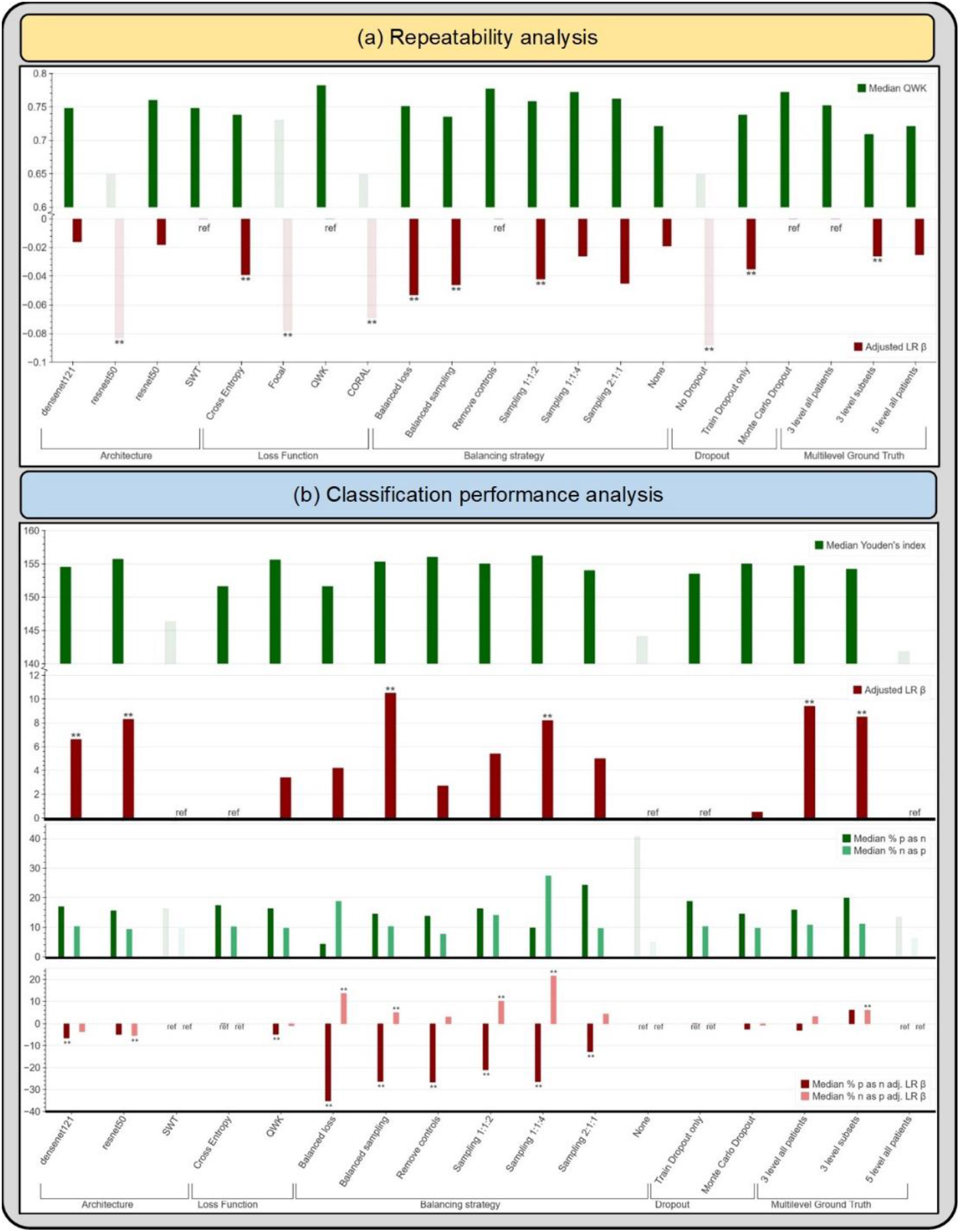
(a) Median quadratic weighted kappa (QWK) and adjusted linear regression (LR) β across the various design choices, as part of the repeatability analysis. (b) Median Youden’s index, median % precancer+ as normal (% p as n) and median % normal as precancer+ (% n as p), with the corresponding adjusted LR β values across the various design choices (after filtering for repeatability), as part of the classification performance analysis. SWT: Swin Transformer; CORAL: CORAL (consistent rank logits) loss, as described in the METHODS section; ref: reference category.

### CLASSIFICATION PERFORMANCE ANALYSIS

Table 3 highlights the summary of the classification performance analysis (Stage II), reporting the median and the interquartile ranges for each of our two key classification metrics: 1. Youden’s index and 2. extreme misclassifications, as well as the adjusted linear regression β for each design choice. Similar to Stage 1, we evaluated the metrics both overall and within each design choice category, dropping the worst performing design choices at this stage in a two-level approach.

**TABLE 3:** Classification performance analysis highlighting Youden’s index (YI) and extreme misclassification statistics – median with interquartile range (IQR) and adjusted linear regression (LR) β values – for design choices within each design choice category for our automated visual evaluation (AVE) classifier, after filtering for repeatability (Table 2). Rows shaded in salmon indicate design choices filtered out at this stage due to poor classification performance (as captured by the Youden’s index). Rows shaded in gray indicate design choices subsequently filtered out due to a combination of poor classification performance (as captured by the rate of extreme misclassifications) and/or practical reasons. SWT: Swin Transformer; ref: reference category.

| Table 3: Classification performance analysis |  |  |  |  |  |  |  |  |  |  |
| --- | --- | --- | --- | --- | --- | --- | --- | --- | --- | --- |
| Design Choice Category | Design Choices | Youden's index (YI) |  |  | Extreme misclassifications |  |  |  |  |  |
|  |  |  |  |  | % precancer+ as normal |  |  | % normal as precancer+ |  |  |
| | | Median (IQR) | | Adjusted LR $\beta$ | Median (IQR) | | Adjusted LR $\beta$ | Median (IQR) | | Adjusted LR $\beta$ |
| Architecture | densenet121 | 154.5 | (151.5 - 156.3) | 6.6** | 17.0 | (10.9 - 23.2) | -6.5** | 10.3 | (6.8 - 13.6) | -3.6 |
|  | resnet50 | 155.7 | (151.7 - 157.9) | 8.3** | 15.6 | (11.6 - 23.9) | -4.9** | 9.3 | (5.7 - 12.2) | -5.4** |
|  | SWT | 146.3 | (134.7 - 148.0) | ref | 16.3 | (13.0 - 56.5) | ref | 9.5 | (4.7 - 14.6) | ref |
| Loss Function | Cross Entropy | 151.6 | (144.1 - 155.7) | ref | 17.4 | (11.2 - 37.3) | ref | 10.2 | (5.3 - 14.5) | ref |
|  | QWK | 155.6 | (153.7 - 157.6) | 3.4 | 16.3 | (11.6 - 21.0) | -4.8** | 9.7 | (7.6 - 11.7) | -0.9 |
| Balancing Strategy | Balanced loss | 151.6 | (142.3 - 154.4) | 4.2 | 4.3 | (3.6 - 5.8) | -35.2** | 18.8 | (10.3 - 23.0) | 13.6** |
|  | Balanced sampling | 155.3 | (153.3 - 157.8) | 10.5** | 14.5 | (13.0 - 18.1) | -26.3** | 10.3 | (8.7 - 11.9) | 4.9** |
|  | Remove controls | 156.0 | (153.5 - 156.9) | 2.7 | 13.8 | (10.9 - 18.1) | -26.6** | 7.7 | (4.2 - 10.3) | 2.9 |
|  | Sampling 1:1:2 | 155.0 | (153.6 - 156.0) | 5.4 | 16.3 | (12.0 - 21.4) | -21.0** | 14.1 | (11.3 - 17.4) | 10.1** |
|  | Sampling 1:1:4 | 156.2 | (151.4 - 158.4) | 8.2** | 9.8 | (6.2 - 14.1) | -26.4** | 27.4 | (15.9 - 38.5) | 21.6** |
|  | Sampling 2:1:1 | 154.0 | (152.9 - 154.5) | 5.0 | 24.3 | (23.2 - 25.0) | -12.7** | 9.6 | (7.4 - 11.4) | 4.2 |
|  | None | 144.1 | (135.2 - 148.9) | ref | 40.6 | (37.0 - 55.8) | ref | 5.0 | (2.3 - 6.6) | ref |
| Dropout | Train Dropout only | 153.5 | (148.8 - 155.7) | ref | 18.8 | (12.3 - 25.4) | ref | 10.3 | (6.7 - 14.1) | ref |
|  | Monte Carlo Dropout | 155.0 | (146.0 - 157.2) | 0.5 | 14.5 | (9.4 - 22.5) | -2.5 | 9.7 | (5.1 - 14.2) | -0.7 |
| Multilevel Ground Truth | 3 level all patients | 154.7 | (151.6 - 156.8) | 9.4** | 15.9 | (10.5 - 23.6) | -3.0 | 10.8 | (6.8 - 15.2) | 3.1 |
|  | 3 level subsets | 154.2 | (153.0 - 156.7) | 8.5** | 19.9 | (18.1 - 23.2) | 6.0 | 11.1 | (9.5 - 13.4) | 5.9** |
|  | 5 level all patients | 141.8 | (135.3 - 151.8) | ref | 13.4 | (10.9 - 50.7) | ref | 6.2 | (4.8 - 9.5) | ref |

In the first level, we looked at the Youden’s index across all design choices and dropped the worst performing choices; this resulted in 3 choices (SWT architecture, no balancing, 5-level ground truth) or 17.6% of the remaining choices being dropped and amounted to dropping choices that had median Youden’s index of <150 (Table 3, shaded in salmon; Fig. 3b, muted bars); this was further supported by other design choices within each design choice category having positive adjusted linear regression β values. In the second level, we considered two factors: 1. median extreme misclassification percentages (% precancer+ as normal and % normal as precancer+); and 2. practical reasons, dropping design choices due to a combination of these two factors. This resulted in three balancing strategies (Sampling 1:1:2, 1:1:4 and 2:1:1) and the “3 level subsets” ground truth mapping, or 28.6% of the remaining design choices being dropped (Table 3, shaded in gray). Weighted sampling by using preassigned label weights per class for the loading sampler (such as 1:1:4) is imprecise since weights are not adjusted relative to the dataset-specific class imbalance; this skews the model in making predictions along the lines of the assigned weights. This can be seen among the sampling strategies dropped: sampling 1:1:4 had a high rate of median % normal predicted as precancer+ (27.4%), while sampling 2:1:1 had a high rate of median % precancer+ predicted as normal (24.3%). The “3 level subsets” ground truth mapping was dropped for practical reasons: it was generated from the 5-level map by omitting the GL and GH labels to attempt to generate further distinction or discontinuity between the three classes (normal, GM, precancer+) during model experimentation. Both the “5-level all patients” and the “3-level subsets” ground-truth mapping are impractical due to the limited clinical data (either HPV, histology and/or cytology) we anticipate having available in the field to generate 5 distinct levels of ground truth, thereby rendering retraining, validation and implementation of these approaches challenging.

### HPV-GROUP COMBINED RISK STRATIFICATION ANALYSIS

Fig. 4 and Table 4 highlight the 10 best performing models that emerge following Stages 1, 2 and 3 of our model selection approach. All 10 models perform similarly among HPV positive women in the full 5-study set, while showing notable differences per study as shown in the NHS subset of the full 5-study set, measured by the combined HPV-AVE AUC. The NHS subset represents women who are closer to a screening population that we would expect in the field when considering deployment of our model, since this is a population-based cohort study (35); hence AUC on the NHS subset represents a truer metric for model comparison. The models in Fig. 4a and Table 4 are in decreasing order of AUC on the HPV positive NHS subset. Fig. 4b plots the ROC curves for each of the top 4 out of the 10 models highlighted in Table 4 and Fig. 4a, highlighting 1. HPV risk-based stratification; 2. model stratification; and 3. combined stratification incorporating both HPV risk and model predicted class.

**FIGURE 4:**
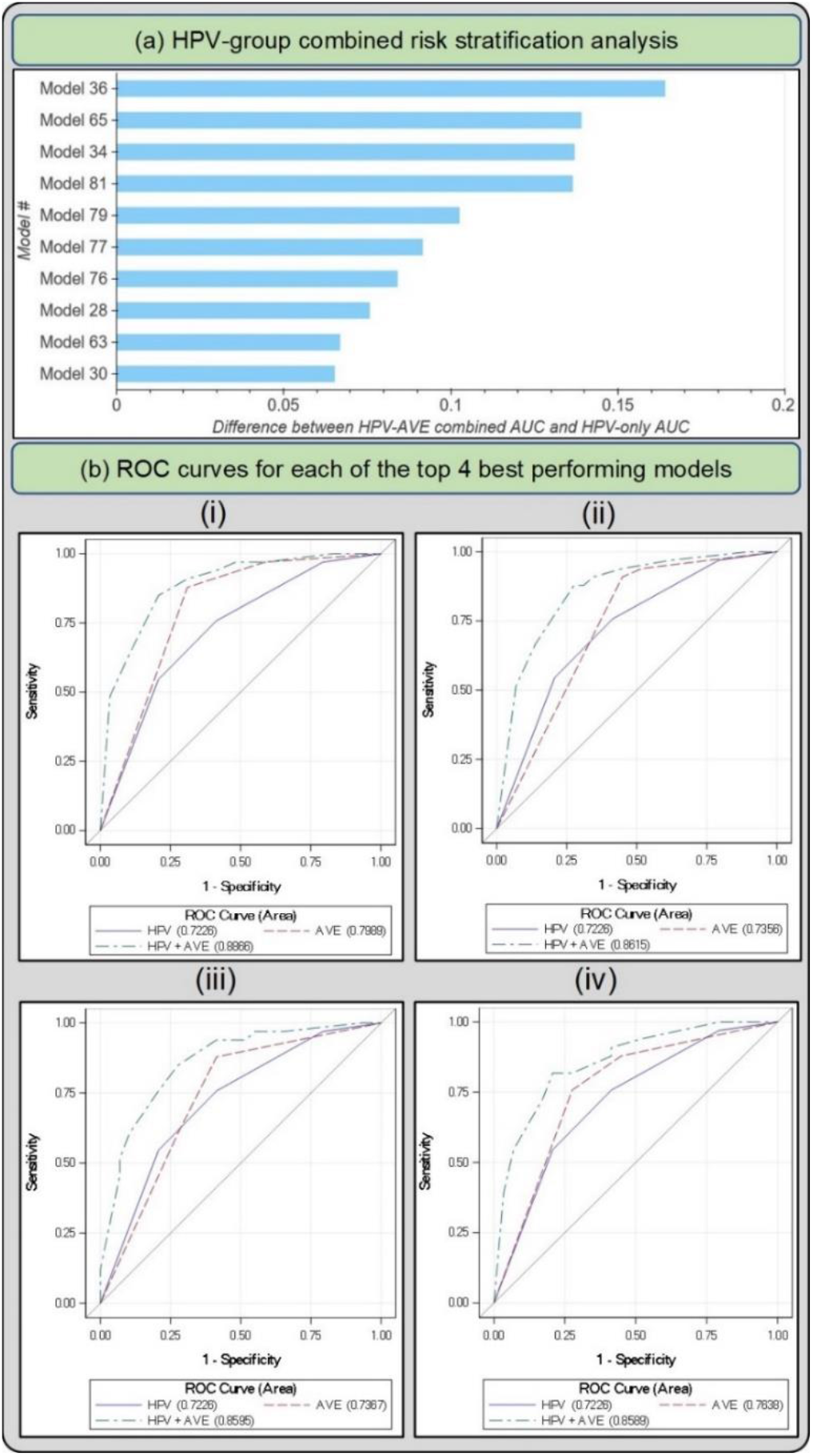
(a) Difference between HPV+AVE combined AUC and HPV-only AUC in the HPV positive NHS subset for top 10 models (b) Receiver operating characteristics (ROC) curves for each of the top 4 best performing models in the HPV positive NHS subset of the full dataset The plotted lines indicate 1. HPV AUC, 2. AVE AUC and 3. combined HPV-AVE AUC, for models (i) 36, (ii) 65, (iii) 34, and (iv) 81. HPV: human papillomavirus; AVE: automated visual evaluation, which refers to the classifier; AUC: area under the ROC curve.

**TABLE 4:**
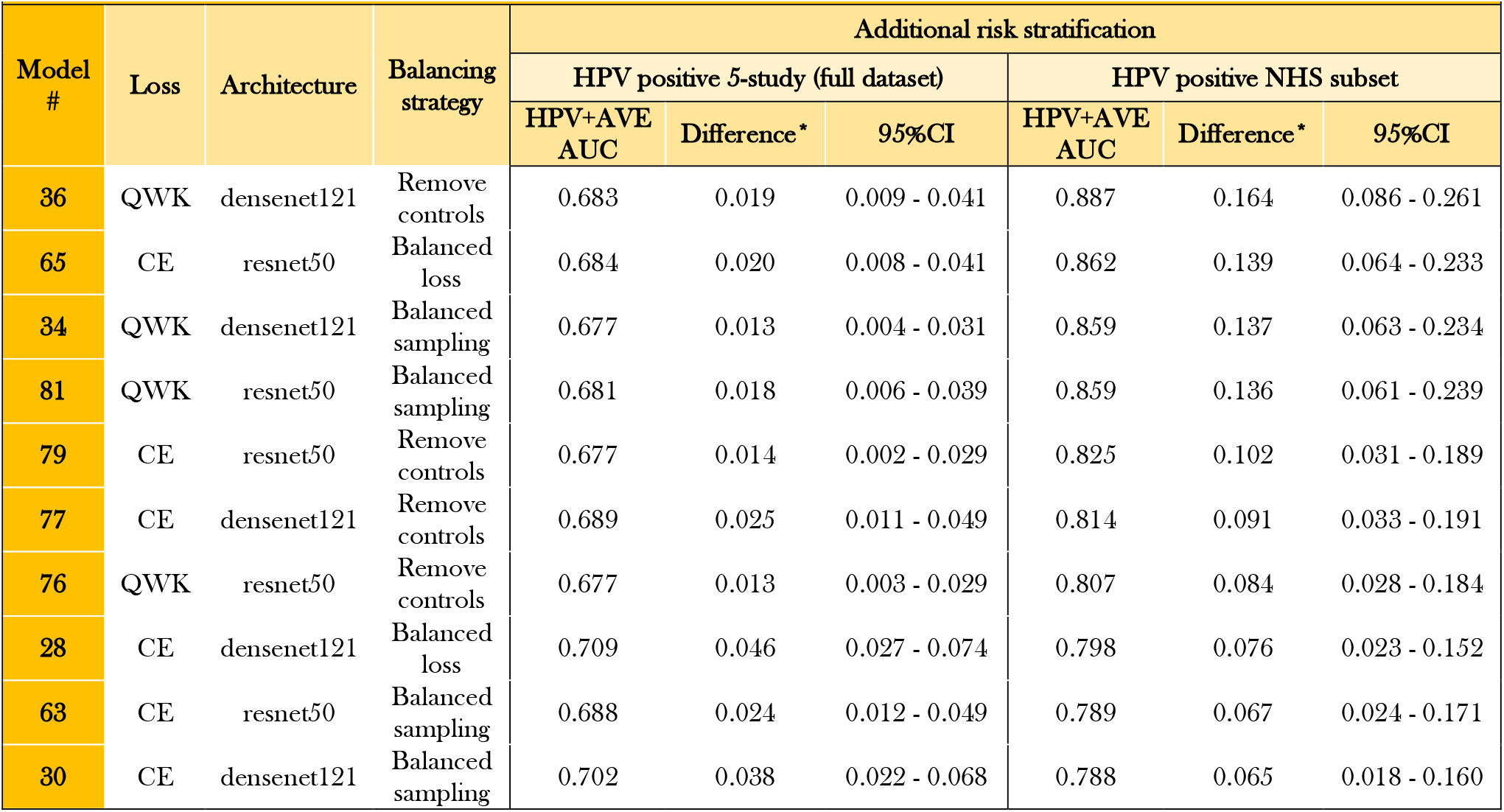
Performance of top individual models following human papillomavirus (HPV) group combined risk stratification (Stage III of model selection) on Test Set 1, within the HPV-positive full-dataset and HPV-positive NHS subset. The models are in decreasing order of area under the receiver operating characteristics (ROC) curve (AUC) on the human papillomavirus (HPV) positive NHS subset of the full dataset. AVE: automated visual evaluation, which refers to the classifier; CI: confidence interval. *Difference = Combined HPV+AVE AUC minus HPV-only AUC.

### CLASSIFICATION AND REPEATABILITY ANALYSIS: TEST SET 2

Fig. 5a and Table 5 highlight the additional classification (1. % precancer+ as normal and 2. % normal as precancer+), and repeatability (1. % 2-class disagreement and 2. QWK) metrics from the predictions of each of the top 10 models on Test Set 2, while Figure 6 takes a deeper look by comparing individual model predictions across 60 images for these top 10 models on Test Set 2. The top 10 models that pass through all stages of our model selection approach utilize the following configurations:

**FIGURE 5:**
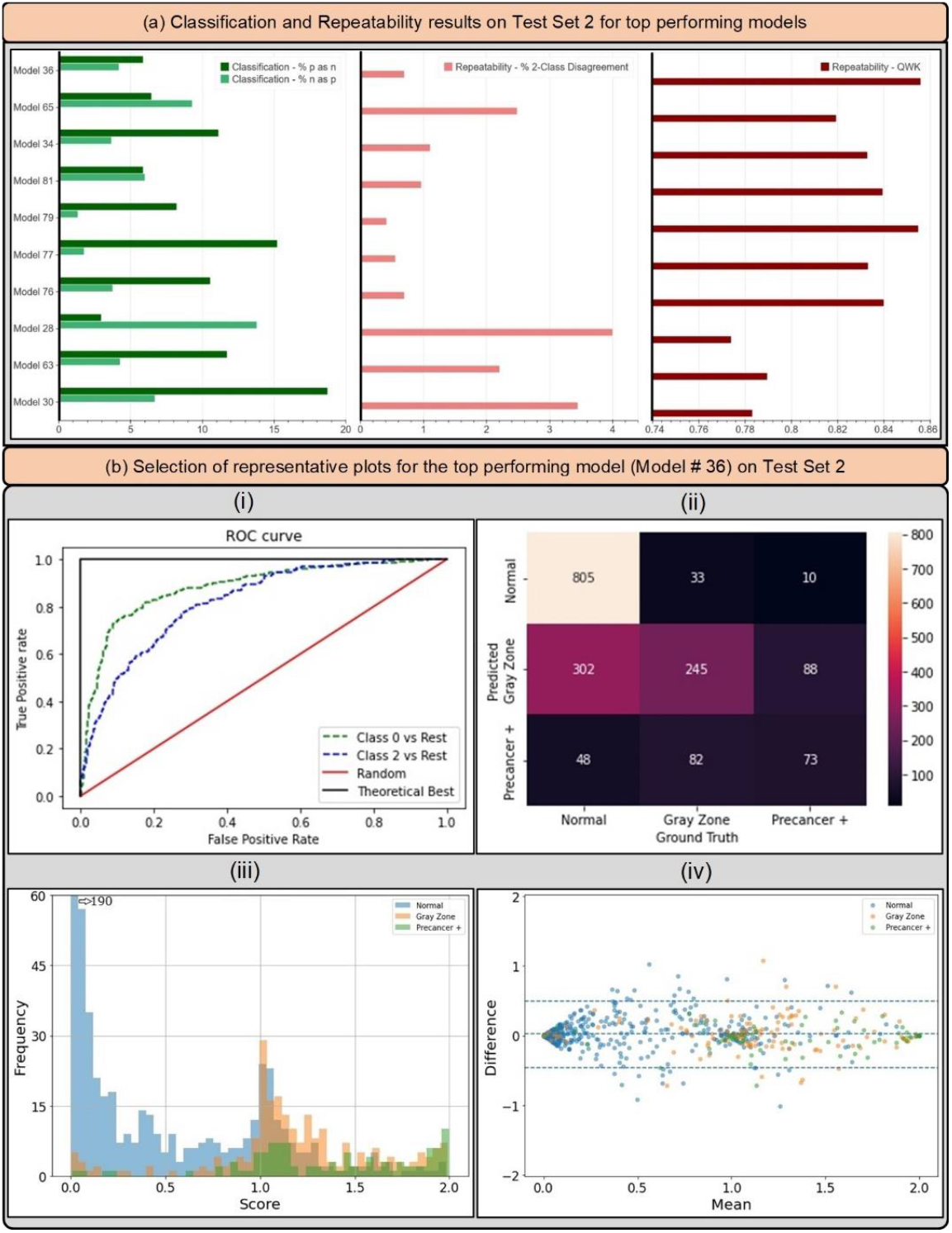
(a) Classification and repeatability results on Test Set 2 for top 10 best performing models, highlighting the % precancer+ as normal (%p as n) and % normal as precancer+ (%n as p) (left), the % 2-class disagreement between image pairs across women (middle), and the quadratic weighted kappa (QWK) values on the discrete class outcomes for paired images across women (right) for each model. (b) Representative plots for the top performing model (# 36) on Test Set 2 - (i) Receiver operating characteristics (ROC) curves for the normal vs rest (Class 0 vs. rest) and precancer+ vs. rest (Class 2 vs. rest) cases, (ii) confusion matrix, (iii) histogram of model predicted continuous *score*, color coded by ground truth, and (iv) Bland Altman plot of model predictions, color coded by ground truth: each point on this plot refers to a single woman, with the y-axis representing the maximum difference in the score across repeat images per woman, and the x-axis plotting the mean of the corresponding score across all repeat images per woman.

**TABLE 5:**
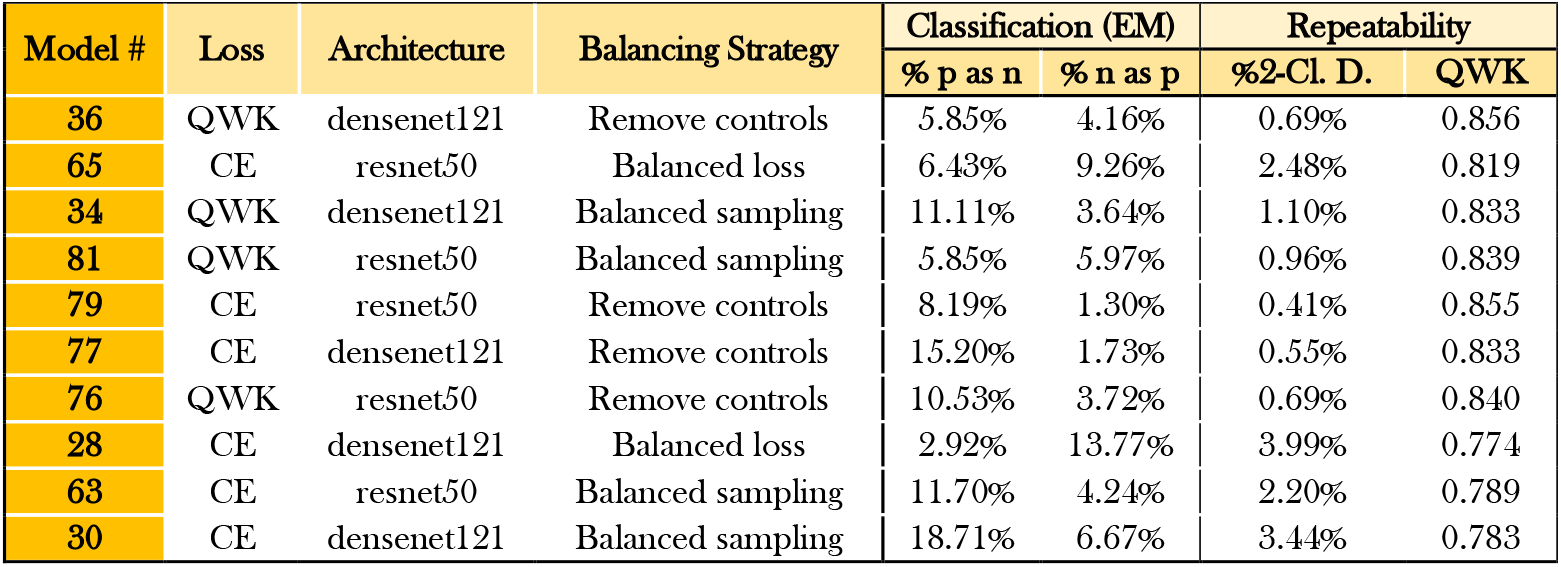
Classification and repeatability results on Test Set 2 for top 10 best performing models, highlighting % precancer+ as normal (% p as n) and % normal as precancer+ (% n as p), the % 2-class disagreement between image pairs across women (% 2-Cl. D.), and the quadratic weighted kappa (QWK) values on the discrete class outcomes for paired images across women, for each model. EM: extreme misclassifications.

| Table 5: Classification and Repeatability results on Test Set 2 for top performing models |  |  |  |  |  |  |  |
| --- | --- | --- | --- | --- | --- | --- | --- |
| Model # | Loss | Architecture | Balancing Strategy | Classification (EM) |  | Repeatability |  |
|  |  |  |  | % p as n | % n as p | %2-Cl. D. | QWK |
| 36 | QWK | densenet121 | Remove controls | 5.85% | 4.16% | 0.69% | 0.856 |
| 65 | CE | resnet50 | Balanced loss | 6.43% | 9.26% | 2.48% | 0.819 |
| 34 | QWK | densenet121 | Balanced sampling | 11.11% | 3.64% | 1.10% | 0.833 |
| 81 | QWK | resnet50 | Balanced sampling | 5.85% | 5.97% | 0.96% | 0.839 |
| 79 | CE | resnet50 | Remove controls | 8.19% | 1.30% | 0.41% | 0.855 |
| 77 | CE | densenet121 | Remove controls | 15.20% | 1.73% | 0.55% | 0.833 |
| 76 | QWK | resnet50 | Remove controls | 10.53% | 3.72% | 0.69% | 0.840 |
| 28 | CE | densenet121 | Balanced loss | 2.92% | 13.77% | 3.99% | 0.774 |
| 63 | CE | resnet50 | Balanced sampling | 11.70% | 4.24% | 2.20% | 0.789 |
| 30 | CE | densenet121 | Balanced sampling | 18.71% | 6.67% | 3.44% | 0.783 |

**FIGURE 6:**
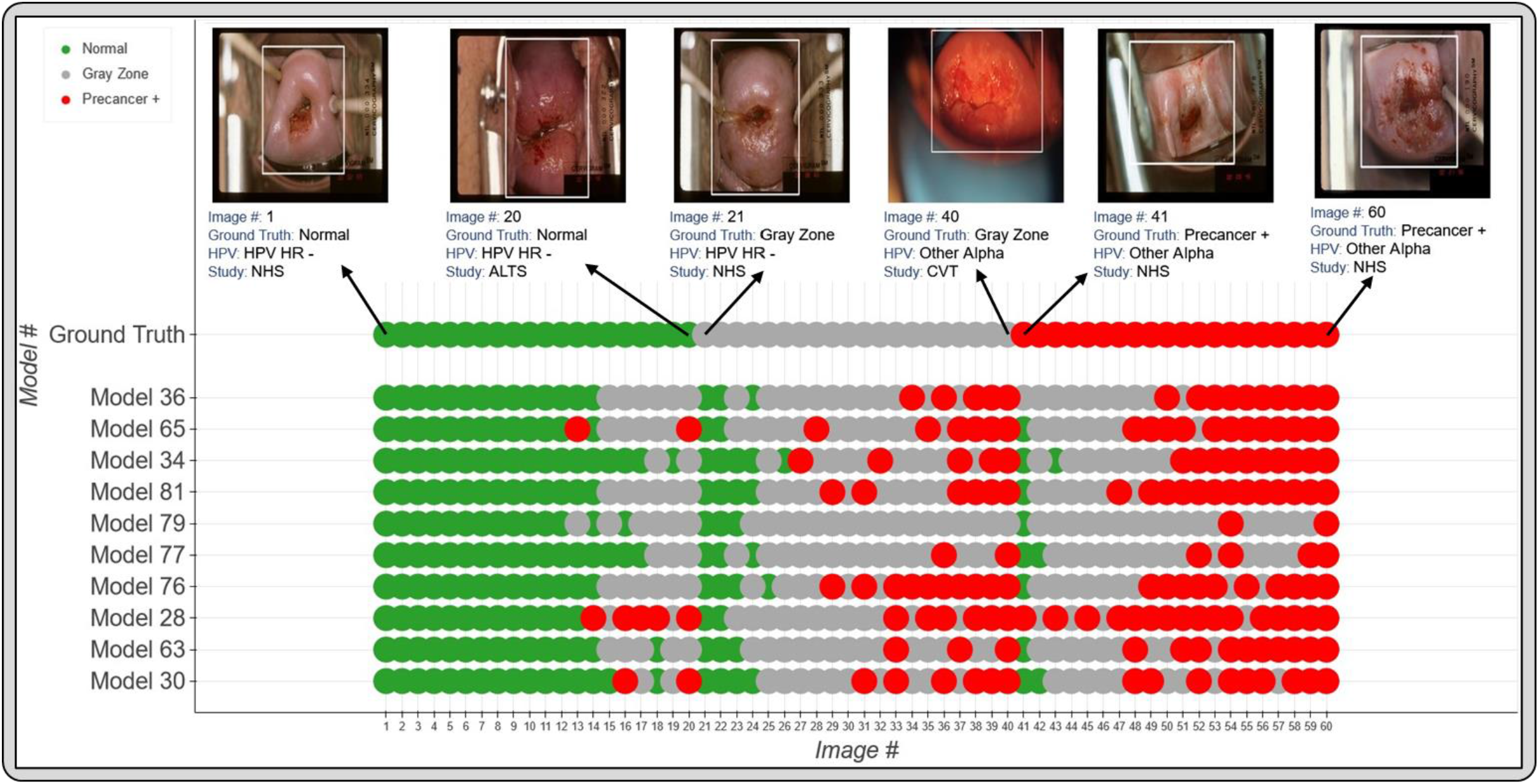
Model level comparison across top-10 best performing models. 60 images were randomly selected (see METHODS: Statistical Analysis section) and arranged in order of increasing mean score within each ground truth class in the top row (labelled “Ground Truth”). The model predicted class for the top 10 models for each of these 60 images is highlighted in the bottom rows, where the images follow the same order as the top row. The color coding in the top row represents ground truth while in the bottom 10 rows represent the model predicted class. Green: Normal, Gray: Gray Zone, and Red: Precancer +, as highlighted in the legend. Each image corresponds to a different woman.

- Architecture: densenet121 or resnet50
- Loss function: quadratic weighted kappa (QWK) or cross-entropy (CE)
- Balancing strategy: remove controls or balanced sampling
- Dropout: Monte-Carlo (MC) dropout (spatial)
- Multi-level ground truth: 3 level all patients (Normal, Gray Zone, Precancer+)
- Model type: multiclass classification

Based on the individual performances of the models in terms of degree of extreme misclassifications and repeatability (Table 5, Fig. 5a) and additional risk stratification (Table 4, Fig. 4), our best performing model (# 36) has the smallest rate of overall extreme misclassifications (5.9% precancer+ as normal, 4.2% normal as precancer+), one of the highest repeatability performance (repeatability QWK = 0.8557, 0.69% 2-class disagreement on repeat images across women), and the highest additional risk stratification in the NHS subset of the full 5-study dataset, our screening population (difference between HPV-AVE combined AUC and HPV AUC= 0.164). Among the top 10 models, model # 36 utilizes the following unique design choices:

- Architecture: densenet121
- Loss function: quadratic weighted kappa (QWK)
- Balancing strategy: remove controls

Fig. 5b highlights key performance metrics of the top ranked model (# 36) on Test Set 2, as captured by the corresponding (i) ROC curves, (ii) confusion matrix, (iii) histogram of the model predicted *score* and (iv) Bland-Altman plot. The ROC curve in (i) demonstrates excellent discrimination of the normal (class 0) and precancer+ (class 2) categories, with corresponding AUROC’s of 0.88 (class 0 vs. rest) and 0.82 (class 2 vs. rest) respectively. This is reinforced by the confusion matrix in (ii), which highlights a total extreme misclassification (extreme off diagonals) rate of only 3.4%, and by the histogram in (iii), which illustrates the strong class separation in model predicted *score*; specifically, (iii) highlights that the model confidently predicts the largest clusters of each of the three ground truth classes correctly as shown by the peaks around *score* 0.0, 1.0 and 2.0. Finally, the Bland-Altman plot in (iv) highlights the model performance in terms of repeatability: each point on this plot refers to a single woman, with the y-axis representing the maximum difference in the *score* across repeat images per woman, and the x-axis plotting the mean of the corresponding *score* across all repeat images per woman. Repeatability is evaluated using the 95% limits of agreement (LoA), highlighted by the blue dotted lines in (iv) on either side of the mean (central blue dotted line); for model # 36, the 95% LoA is quite narrow, with most points clustered around 0 on the y-axis suggesting that *score* values of the model on repeat images taken on the same visit for each woman are quite similar; here, the 95% LoA adjusted for the number of classes and presented as a fraction of the possible value range is 0.240 (±0.038).

Fig. 6 reinforces the validity of our approach for model selection and optimization by providing a detailed comparison of model performance at the individual image level, with the top models performing desirably with respect to the clinical problem we are aiming to address. Incorporation of a gray zone class, together with MC dropout and loss functions that penalize misclassifications between the extreme classes ensures that we deal with ambiguity with cases at the class boundaries. For instance, among these randomly selected 60 images, the best performing model (# 36) has the lowest rate of extreme misclassifications (none), while predicting a wide enough gray zone that adequately encapsulates the clinical ambiguity with uncertain cases: these are cases for which even clinically trained colposcopists and gynecologic oncologists would find determination of precancer+ status challenging.

## DISCUSSION

Despite the advancements made by AI in clinical classification tasks, key concerns hindering model deployment from bench to clinical practice include model reliability and clinical translatability. An incorrect, unreliable, or unrepeatable model prediction has the potential to lead to a cascade of clinical actions that might jeopardize the health and safety of a patient. Therefore, it is essential that models designed with the goal of clinical deployment be specifically optimized for improved repeatability and clinical translation.

Our work addresses these concerns of reliability and clinical translatability. We optimize our model selection approach with improved repeatability as the primary stage (Stage I) of our selection criterion – ensuring that only design choices that produce repeatable, reliable predictions across multiple images from the same woman’s visit, are passed through to the next stage of evaluation for classification performance. Our work builds on prior work highlighting improvements in repeatability of model predictions made by certain design choices (36,37). Our work also stands out among the paucity of current approaches that have utilized AI and DL for cervical screening (21–24); as aforementioned, these are largely plagued by overfitting and no consideration of repeatability. The dearth of work investigating repeatability of AI models designed for clinical translation in the current DL and medical image classification literature has meant that no rigorous study, to the best of our knowledge, has employed repeatability as a model selection criterion. We posit that our work could motivate further efforts to include repeatability as a key criterion for clinical AI model design.

Subsequent design choices of our work are optimized to improve clinical translatability. Prior work (21–24) has shown us that while binary classifiers for cervical image-based cervical precancer+ detection can achieve competitive performance in a given internal seed dataset, they translate poorly when tested in different settings; uncertain cases can be misclassified, and predictions tend to oscillate between the two classes. This oscillation phenomenon could prevent a precancer+ woman from accessing further evaluation (i.e., false negative) or direct a normal woman through unnecessary, potentially invasive tests (i.e., false positive). False negatives are especially problematic in LMIC where screening is limited and represent a missed opportunity to detect and treat precancer via excisional, ablative, or surgical methods, in order to avert cervical cancer (13,38). By incorporating a multi-class approach and a loss function that heavily penalizes extreme misclassifications, we improve reliability of the model-predicted normal and precancer+ categories, and further ensure that women ascribed to the intermediate classes are recommended for additional clinical evaluation.

Finally, our choice of incorporating HPV genotyping together with model predictions and assessing model performance based on the ability to further stratify precancer+ risk associated with each of the four groups of high-risk HPV types, is very relevant for cervical screening. Recent work has shown that the presence of clinical variables as additional inputs to a neural network can both enhance model performance and lend interpretability to the value of these variables for clinical decision making (5,39,40). Incorporating relevant clinical data and prognostic variables is an approach that, we believe, should become standard for cancer classifier design, and in particular for neoplasms with well-known clinical causative agents.

Our prior work has informed us that the HPV positive women in the NHS subset better represent a typical screening population: specifically, the NHS subset represents women who tested HPV-positive in any given population with an intermediate HPV prevalence (35). The other 4 subsets within the full 5-study dataset comprise of women referred from HPV-based/cytology-based referral clinics: this represents a colposcopy population, which has a higher disease prevalence. We optimize each stage (I, II and III) of our model selection approach on the full 5-study dataset to better capture the variability in cervical appearance on imaging. At the end of this selection, we find that our top models do not perform meaningfully differently among HPV positive women in the full 5-study dataset, highlighted by similar HPV-AVE AUC values across the models in the “HPV positive 5 study” column on Table 4. For the final selection of the top candidates, given our goal of using AVE as a triage tool for HPV positive women in a screening setting, we therefore narrow our focus to the combined HPV-AVE AUC in the NHS HPV positive subset (“HPV positive NHS” column on Table 4; Fig. 4) for each model on Test Set 1 and confirm performance of the top candidates on Test Set 2 (Table 5, Fig. 5a).

Despite the multi-institutional, multi-device and multi-population nature of our final, collated dataset; the use of multiple held-aside test sets; and the exhaustive search space utilized for our algorithm choices, our work may be limited by sparse external validation. Forthcoming work will evaluate our model selection choices on several additional external datasets, assessing out-of-the-box performance as well as various transfer learning, retraining and generalization approaches. Future work will additionally optimize our final model choice for use on edge devices, thereby promoting deployability and translation in LMIC.

In this work, we utilized a large, multi-institutional, multi-device and multi-population dataset of 9,462 women (17,013 images) as a seed and implemented a comprehensive model selection approach to generate a diagnostic classifier, termed AVE, able to classify images of the cervix into “normal”, “gray zone” and “precancer+” categories. Our model selection approach investigates various choices of model architecture, loss function, balancing strategy, dropout, and ground truth mapping, and optimizes for 1. improved repeatability; 2. classification performance; and 3. high-risk HPV-type-group combined risk-stratification. Our best performing model uniquely 1. alleviates overfitting by incorporating spatial MC dropout to regularize the learning process; 2. achieves strong repeatability of predicted class across repeat images from the same woman; 3. addresses rater and model uncertainty with ambiguous cases by utilizing a three-level ground truth and QWK as the loss function to penalize extreme (between boundary class) misclassifications; and 4. achieves a strong additional risk-stratification when combined with the corresponding HPV type group within our screening population of interest. While our initial goal is to implement AVE primarily to triage HPV positive women in a screening setting, we expect our approach and selected model to also provide reliable predictions both for images obtained in the colposcopy setting, as well as in the absence of HPV results. Our model selection approach is generalizable to other clinical domains as well: we hope for our work to foster additional, carefully designed studies that focus on alleviating overfitting and improving reliability of model predictions, in addition to optimizing for improved classification performance, when deciding to use an AI approach for a given clinical task.

## METHODS

### OVERVIEW

This study set out to systematically compare the impact of multiple design choices on the ability of a deep neural network (DNN) to classify cervical images into delineated cervical cancer risk categories. We combined images of the cervix from five studies (Supp. Table 1) into a large convenience sample for analysis. We subsequently labelled the images into three distinct multi-level ground truth labelling approaches: 1. a 5-level map, which included normal, gray-low (GL), gray-middle (GM), gray-high (GH), and precancer+ (termed “5 level all patients”); 2. a 3-level map which combined the intermediate three labels (GL, GM, GH) into one single gray zone (termed “3 level all patients”); and 3. an additional 3-level map which excluded the GL and GH labels, and considered only the normal, GM and precancer+ labels (termed “3 level subsets”). The choice of multi-level ground truth labelling for model selection was motivated by our previous work and intuition revealing the failure of binary models, as well as our specific clinical use case. Table 1 highlights the population level and dataset level characteristics for our final, collated dataset used for training and evaluation, highlighting the distribution of histology, cytology, HPV types, population-level study, age, and number of images per patient within each of the five ground truth classes.

We subsequently identified four key design decision categories that were systematically implemented, intersected, and compared. These included: model architecture, loss function, balancing strategy, and implementation of dropout, as highlighted in Fig. 1. The choice of balancing strategy for a particular model determined the ratios of randomly chosen train and validation sets used during training. We subsequently trained multiple classifiers using combinations of these design choices and generated predictions on a common test set (“Test Set 1”) which allowed for comparison and ranking of approaches based on repeatability, classification performance, and HPV type-group combined risk stratification. Finally, we confirmed the performance of the top models on a second test set (“Test Set 2”) to mitigate the impact of chance on the best performing approaches.

### DATASET

#### Included Studies

Cervical images used in this analysis were collected from five separate study populations labelled NHS, ALTS, CVT, Biop and D Biop (Table 1; Fig. 1). Detailed descriptions for each study can be found in the supplementary methods section. The final dataset was collated into a large convenience sample comprising of a total of 17,013 images from 9,462 women.

#### Analysis population

The convenience sample was split using random sampling into four sets for use in the evaluation of algorithm parameters. For the initial splits, women were randomly selected into either training, validation, or test (“Test Set 1”), at a rate of 60%, 10%, and 20% respectively. An additional hold-back test set (“Test Set 2”) of 10% of the total women was selected and used to confirm the findings of the best models from Test Set 1. All subsets maintained the same study and ground truth proportions as the full set (Table 1, Supp. Table 2). All images associated with the selected visit for each woman were included in the set for which the woman was selected; 7359 women (77.8%) had ≥ 2 images. For a woman identified as precancer or worse (precancer+), the visit at or directly preceding the diagnosis was selected, for women identified as any of the gray zone categories (GL, GM, GH), the visit associated with the abnormality was selected, and for a woman identified as normal, a study visit, if there were more than one, was randomly selected for inclusion.

#### Disease endpoint definitions

Ground truth classification in all studies was based on a combination of histology, cytology, and HPV status with emphasis on strictly defining the highest and lowest categories while pushing marginal results into the middle categories. When referral colposcopy lacked cytology or HPV testing the results from the preceding referral screening visit were used. Ground truth classification was generally consistent across studies; however, the multiple cytology results available in NHS allowed for slightly different classifications. In all studies, histologically confirmed cancer, cervical intraepithelial neoplasia (CIN) 3, or adenocarcinoma in situ (AIS) was considered as precancer+ regardless of referral cytology or HPV, while oncogenic HPV-positive-CIN2 was also considered as precancer+. In NHS, women with 2 or more high grade squamous intraepithelial lesion (HSIL) cytology results that tested positive for HPV 16 were classified as precancer+. In all studies, images identified as atypical squamous cells of undetermined significance (ASCUS) or negative for intraepithelial lesion or malignancy (NILM) with negative oncogenic HPV, or as NILM with missing HPV test were labelled as normal. All other combinations were labelled as equivocal called gray zone, with finer distinctions made for the five-level ground truth classification, splitting the gray zone further into GH, GM, and GL based on specific combinations of cytology and HPV (Supp. Table 1).

#### Ethics

All study participants signed a written informed consent prior to enrollment and sample collection. All five studies were reviewed and approved by multiple Institutional Review Boards including those of the National Cancer Institute (NCI), National Institutes of Health (NIH) and within the institution/country where the study was conducted.

### MODEL

#### Algorithm Design

A compendium of models were trained using a combination of different architectures, model types, loss functions, and balancing strategies. All models were trained for 75 epochs with a batch size of 8 and a learning rate of 10^−5^. The model with the highest summed normal and precancer area under the Receiver Operating Characteristics (ROC) curve (AUC) on the validation set was selected as the best model during training. Before training, all images were cropped with bounding boxes generated from a YOLOv5 (41) model trained for cervix detection, resized to 256&256 pixels, and scaled to intensity values from 0 to 1. During training, affine transformations were applied to the image for data augmentation.

The following popular classification architectures were selected based on literature review and preliminary experiments indicating acceptable baseline performance: ResNet50 (42), ResNest50 (43), DenseNet121 (44), and Swin Transformer (45).

Four different loss functions were evaluated, three for classification models and one for ordinal models. For the classification models, we trained with standard cross entropy (CE), focal (FOC, Equation 1) (46), and quadratic weighted kappa (QWK, Equation 2) (47) loss functions, while all ordinal models leveraged the CORAL loss (Equation 3) (48). QWK is based on Cohen’s Kappa coefficient; unlike unweighted kappa, QWK considers the degree of disagreement between ground truth labels and model predictions and penalizes misclassifications quadratically. Relevant equations are highlighted below:

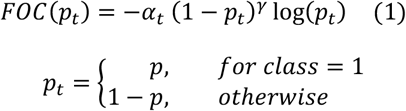

Here, *α*_*t*_ is a weighting factor used to address class imbalance, also present in standard cross-entropy loss implementations, *γ* ≥ 0 is a tunable focusing parameter and *p*_*t*_ is the predicted probability of the ground truth class. We used values of *α*_*t*_ = 0.25 and *γ* = 2, as reported and optimized in previous work (46). Preliminary experiments were also conducted, iterating across *α*_*t*_ = 0.25, 1, *and* inverse class frequency as well as iterating across *γ* = 1.5, 2, 3 *and* 4, before arriving at the optimal choices of *α*_*t*_ = 0.25 and *γ* = 2.

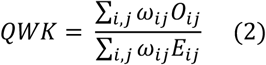

Here, *ω* is the weight matrix for quadratic penalization for every pair 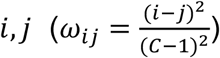, **C** is the number of classes, **O** is the confusion matrix represented by the matrix multiplication between the true value and prediction vectors, and E is the outer product between the true value and prediction vectors.

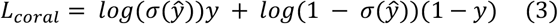

Here σ is the sigmoid function, ŷ is the model’s output, and y is the level-encoded ground truth.

Three balancing strategies were evaluated to deal with the dataset’s class imbalance: weighting the loss function, modifying the loading sampler, and rebalancing the training and validation sets. These strategies were only applied during the training process and were compared against training without balancing. To emphasize the least frequent labels, one approach was to apply weights to the loss function in proportion to the inverse of the occurrence of each class label. A second approach was to reweight the loading sampler to present images associated with each label equally as well as with specific weights – 2:1:1, 1:1:2, or 1:1:4 (Normal : Gray Zone : Precancer+). The final balancing strategy, henceforth termed “remove controls”, involved randomly removing “normal” (class 0) women from the training and validation sets and reallocating them to Test Set 1, in order to better rebalance the training and validation set labels; in this approach, a total of 2383 women (4555 images) from the initial train set, and 410 women (780 images) from the initial validation set were reallocated to the test set. The final class balance in the train and validation sets for the “remove controls” balancing strategy amounted to ∼40% normal : 40% gray zone (including GL, GM, and GH) : 20% precancer+ (Supp. Table 3).

Finally, we evaluated multiple approaches to dropping layers during training to alleviate overfitting and regularize the learning process by randomly removing neural connections from the model (49). Spatial dropout drops entire feature maps during training: a rate of 0.1 was applied after each dense layer for the DenseNet models, and after each residual block for the ResNet and ReNest models. The Swin Transformer models were used as implemented in (45). Monte Carlo (MC) dropout was additionally implemented, which can be thought of as a Bayesian approximation (50) generated by enabling dropout during inference and averaging 50 MC samples. MC models in this work refer to models trained using dropout combined with the inference prediction derived from the 50 forward passes.

### Statistical analysis

Our model selection approach (Fig. 2) consisted of three stages, each utilizing model predictions from Test Set 1. After selection of the 10 best models following stage III, we further evaluated their performance in Test Set 2 to confirm results from Test Set 1.

In Stage I of our model selection approach, we evaluated models based on their ability to classify pairs of cervical images reliably and repeatedly, termed the repeatability analysis. We calculated the QWK values on the discrete class outcomes for paired images from the same woman and visit for all models, calculating the mean, median, and inter-quartile range of the QWK for each design choice. We subsequently ran an adjusted multivariate linear regression of the median QWK vs. the various design choice categories and computed the β values and corresponding p-values for each design choice, holding the design choice with the highest median QWK within each design choice category as reference. This allowed us to gauge the relative impacts from the various design choices within each of the model architecture, loss function, balancing strategy, dropout, and ground truth categories.

In Stage II of our approach, we evaluated classification performance based on two key metrics: 1. Youden’s index, which captures the overall sensitivity and specificity, and 2. the degree of extreme misclassifications; this is termed the classification performance analysis. We computed both sets of metrics for each of the design choices within each design choice category. Our choice to include misclassification of the extreme classes (i.e., precancer+ classified as normal or extreme false negative, and normal classified as precancer+ or extreme false positive) as metrics was motivated by the importance of these metrics for triage tests (51). Similar to the repeatability analysis, we calculated the mean, median, and interquartile ranges for these metrics, as well as conducted separate multivariate linear regressions of each of the three median statistics vs. the various design choices categories; we computed the β values and corresponding p-values holding the design choice with the lowest median Youden’s index within each design choice category as reference. This allowed for comparison across design choices overall and within each design choice category.

In Stage III of our model selection approach, we selected the best individual models determined by their ability to further stratify the risk of precancer associated with each of four groups of oncogenic high-risk HPV-types. HPV screening is known to have an extremely high negative predictive value (52,53), and our approach was motivated by the goal of designing an algorithm to triage HPV positive primary screening. The HPV types were grouped hierarchically in four groupings, in order of decreasing risk (54): 1. HPV 16; 2. HPV 18 or 45; 3. HPV 31, 33, 35, 52, 58; and 4. HPV 39, 51, 56, 59, 68. In order to assess the ability of a model to further stratify HPV associated risk, we ran logistic regression models on a binary precancer+ vs. <precancer variable. These models were adjusted for hierarchical HPV type group and the model predicted class. We subsequently calculated the difference in AUC between the model adjusted for both predicted class and HPV type group and the model adjusted only for HPV type group and highlighted the 10 models with the best additional stratification (Table 4, Fig. 4).

Finally, we computed additional classification performance metrics (1. % precancer+ as normal; and 2. % normal as precancer+), and repeatability metrics (1. the % 2-class disagreement between image pairs; and 2. QWK values, on the discrete class outcomes for paired images across woman) for each of the top 10 models on Test Set 2 (Table 5, Fig. 5), in order to further confirm the performance of these models. Additionally, to aid better visualization of predictions at the individual model level, we generated Figure 6 which compares model predictions across 60 images for each of the top 10 models. To generate this comparison, we first summarized each model’s output as a continuous severity *score*. Specifically, we utilized the ordinality of our problem and defined the continuous severity *score* as a weighted average using softmax probability of each class as described in Equation 3, where *k* is the number of classes and *p*_*i*_ the softmax probability of class *i*.

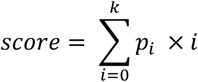

Put another way, the *score* is equivalent to the expected value of a random variable that takes values equal to the class labels, and the probabilities are the model’s softmax probability at index *i* corresponding to class label *i*. For a three-class model, the values lie in the range 0 to 2. We next computed the average of the *score* for each image across all 10 models and arranged the images in order of increasing *score* within each class. From this *score*-ordered list, we randomly selected 20 images per class, maintaining the distribution of mean scores within each class, and arranged the images in order of increasing average *score* within each class in the top row of Fig. 6, color coded by ground truth. We subsequently compared the predicted class across the 10 models for each of these 60 images (bottom 10 rows of Figure 5), maintaining the images in the same order as the ground truth row and color-coded by model predicted class. This enabled us to gain a deeper insight and to compare model performance at the individual image level.

## Data Availability

All data produced in the present work are contained in the manuscript.

## SUPPLEMENTARY INFORMATION

### SUPPLEMENT SECTION 1: SUPPLEMENTARY METHODS

#### (A) INDIVIDUAL DATASET DESCRIPTIONS

##### (i) Natural History Study (NHS)

The Natural History Study (NHS) is a population-based prospective study carried out in Guanacaste Costa Rica between 1993 and 2000 (35). This cohort enrolled women followed in either an active cohort with visits every 6-12 months or a passive cohort screened once during follow-up between 5-7 years after enrollment. Screening visits included collection of specimens for cytology, human papillomavirus (HPV) testing, and digital images, while histology was collected among women with abnormal colposcopic evaluation. Cytology was assessed via both conventional and liquid-based methods as well as a first-generation automated approach. HPV testing by MY09/MY11 polymerase chain reaction (PCR) consensus primers was performed on samples collected by Dacron swabs, however, these results were not used for colposcopy referral during the study. Two cervical images per visit were collected at each screening visit using a Cervigram cerviscope, which were later digitized and compressed for storage (55).

##### (ii) ASCUS/LSIL Triage Study for Cervical Cancer (ALTS)

The ASCUS/LSIL Triage Study for Cervical Cancer (ALTS) is a multi-center randomized trial of US women conducted between 1996 and 2000. This study enrolled women attending colposcopy clinics with referral cytology of either atypical squamous cells of undetermined significance (ASCUS) or low-grade squamous intraepithelial lesion (LSIL). Women were followed for 2 years with screening visits every 6 months. Screening visit specimen collection included two cervical specimens, one for liquid-based cytology and one for HPV testing, as well as cervical images. Referral to colposcopy and histologic sampling varied by study visit, including enrollment referral following the referral cytology result as well as the randomized HPV result, referral from follow-up visit due to high-grade squamous intraepithelial lesion (HSIL) cytology, and exit colposcopy for all women. Type-specific HPV results were not used for patient management (56). Cytologic diagnosis were based on ThinPrep slides created from cytobrush collected exfoliated cells eluted into PreservCyt-media specimens, with both clinical and quality control (QC) evaluations performed. HPV typing was performed by PCR on specimens collected in PreservCyt. A cerviscope was used to collect two images per screening visit and were later converted to a digital format in the same process used for NHS images.

##### (iii) Costa Rica Vaccine Trial (CVT)

The CVT study is a double-blind, controlled, randomized, phase III study of the efficacy of an HPV16/18 virus-like particle (VLP) vaccine in the prevention of advanced cervical intraepithelial neoplasia (cervical intraepithelial neoplasia (CIN) 2, CIN3, adenocarcinoma in situ (AIS) and invasive cervical cancer) associated with HPV 16 or HPV 18 cervical infection in healthy young adult women in Costa Rica, Guanacaste, and parts of the Puntarenas provinces (57). Women were randomized to either the HPV16/18 or control group and followed up for 4 years as part of this study. Images were collected from women who were only referred for colposcopic evaluation, who remained at colposcopy until they had two consecutive results within normal limits. Images were acquired using a Nikon digital single-lens reflex (DSLR) camera with a beam splitter of colposcopy imaging and were subsequently collected using a boundary marking tool.

##### (iv) Biopsy study (Biop)

The Biopsy Study (Biop) was a population-based study of women referred to colposcopy for abnormal cervical cancer screening results conducted at the University of Oklahoma Health Sciences Center (OUHSC) from February 2009 to August 2011, designed with the goal of utilizing biopsies to improve detection of cervical precancer. HPV testing was conducted via the LINEAR ARRAY® multiplexed PCR-based assay. Histologic interpretation of biopsy and LEEP specimens was conducted using CIN terminologies. All women enrolled in the study had a colposcopy performed and at least one biopsy. Images were acquired using a Nikon DSLR camera with a beam splitter of colposcopy imaging and were subsequently annotated and collected using the boundary marking tool (59).

##### (v) Biopsy Study – Europe (D Biop)

Fifth, we used data and images from a European study (D Biop) designed to investigate high-risk HPV genotypes in women with histologic CIN2/3 referred on the basis of abnormal cytology. HPV typing was done on cytology and CIN2/3 biopsies. If the whole-tissue section of the biopsy was positive for multiple high-risk HPV types, LCM-PCR was performed. Images were acquired using a DSLR camera (60).

## SUPPLEMENT SECTION 2: SUPPLEMENTARY TABLES AND FIGURES

**Supplementary Table 1.**
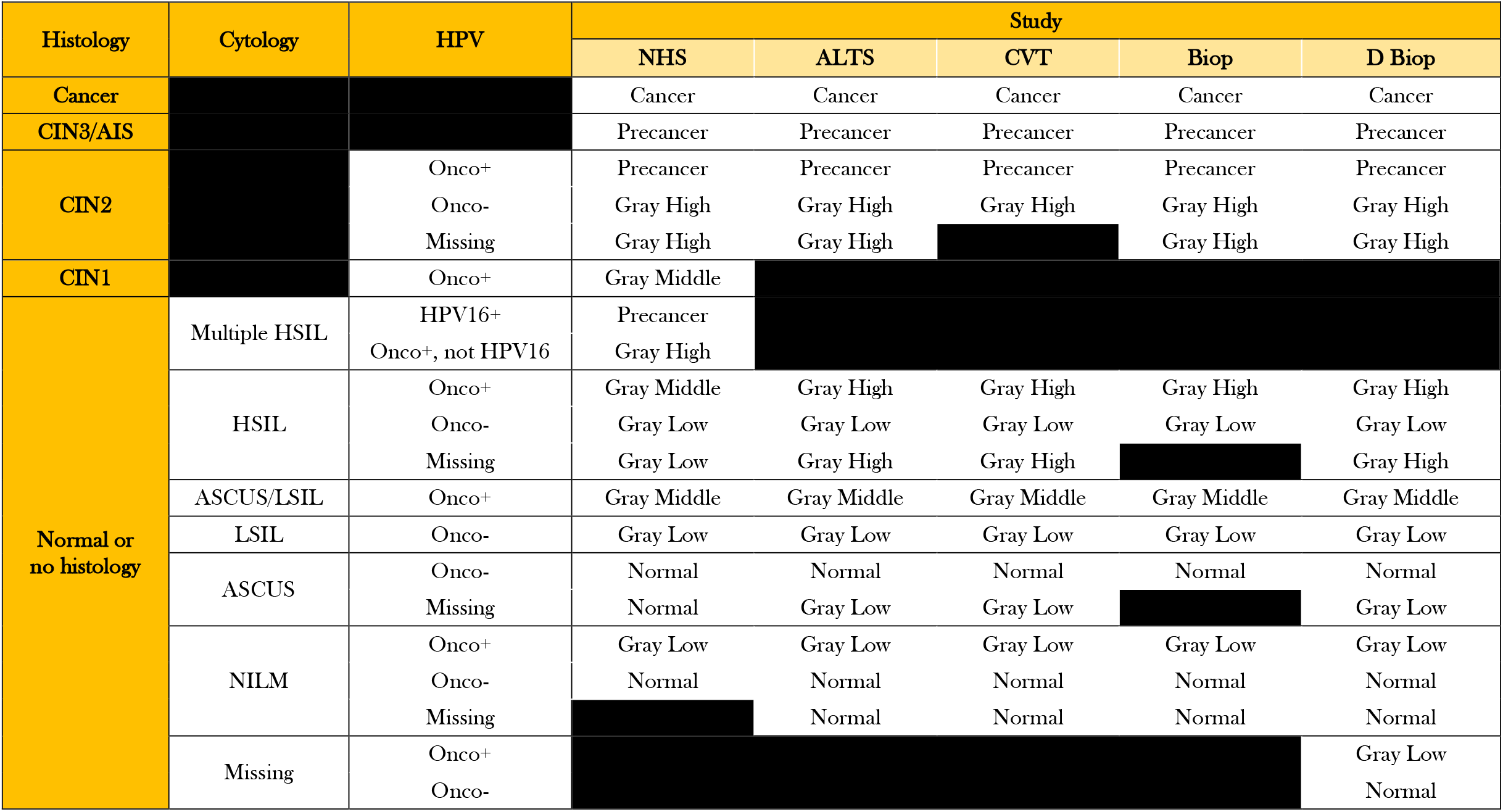
Detailed breakdown of ground truth definitions by study.

**Supplementary Table 2:**
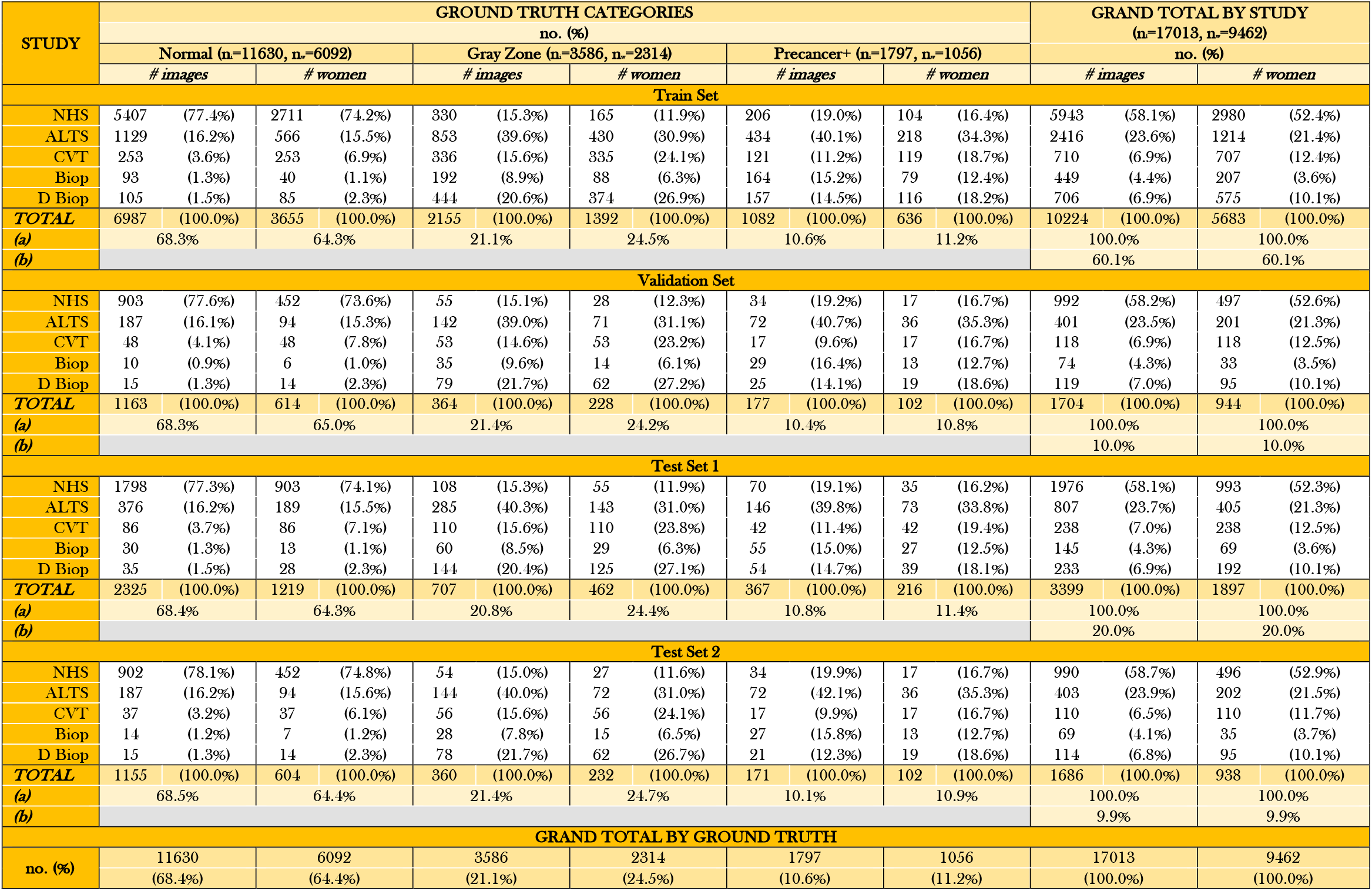
Detailed breakdown of full 5-study dataset by set (train, validation, test 1, test 2), study and ground truth. n_i_=total # images; n_w_=total # women; (a) Ground truth ratios (by images or women) within each set (train/validation/test 1/test 2) = Total # (images or women) in the ground truth category of set ÷ Total # (images or women) in the set; (b) Proportion of total (images or women) in each set (train/validation/test 1/test 2) = Total # (images or women) in the set ÷ Total # (images or women) in the full dataset.

**Supplementary Table 3:**
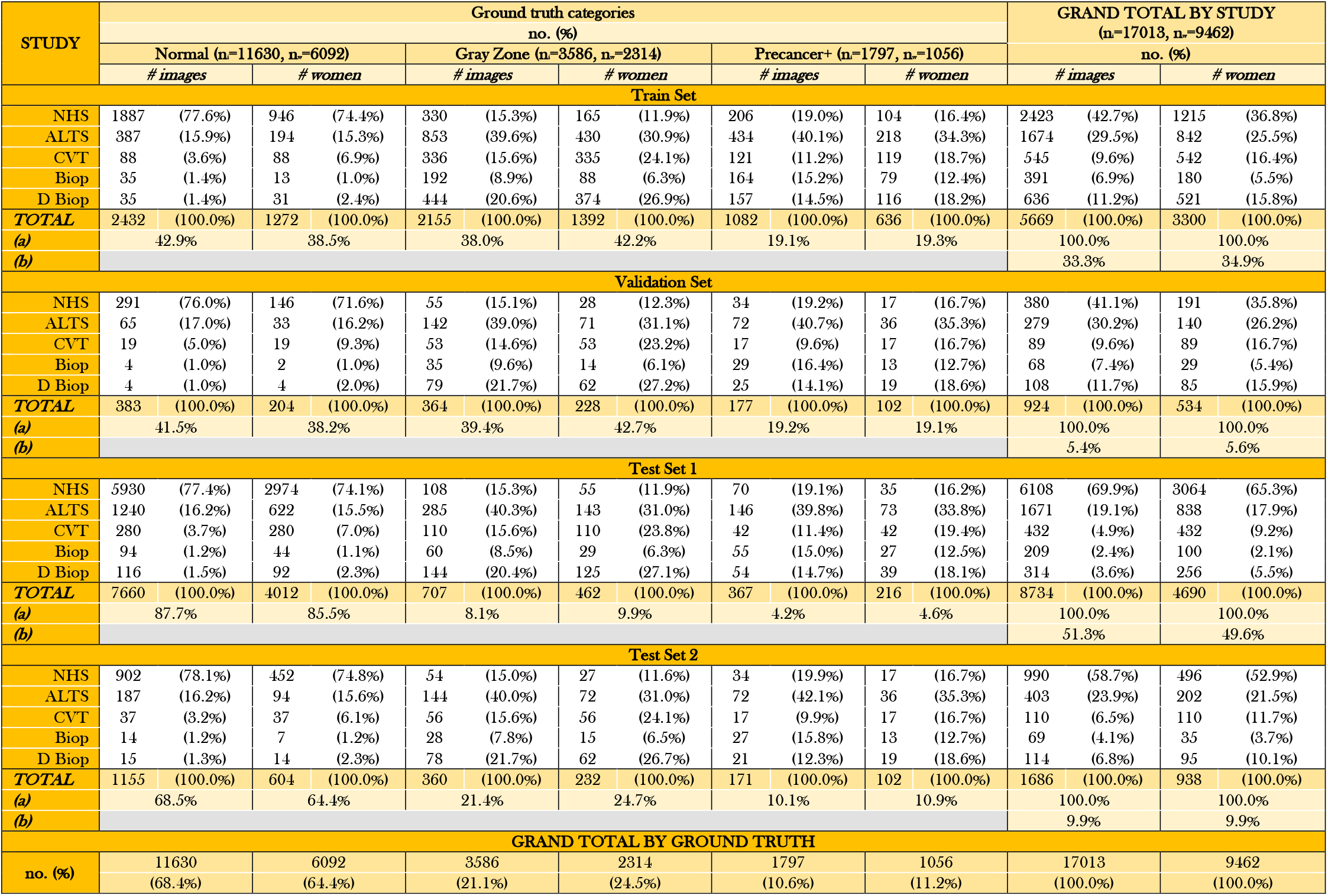
Detailed breakdown of rebalanced dataset after “remove controls” balancing by set (train, validation, test 1, test 2), study and ground truth. n_i_=total # images; n_w_=total # women; (a) Ground truth ratios (by images or women) within each set (train/validation/test 1/test 2) = Total # (images or women) in the ground truth category of set ÷ Total # (images or women) in the set; (b) Proportion of total (images or women) in each set (train/validation/test 1/test 2) = Total # (images or women) in the set ÷ Total # (images or women) in the full dataset.

| Supplementary Table 3: Detailed breakdown of rebalanced dataset after applying “remove controls” balancing, by set ( train, validation, test 1 or test 2), study and ground truth |  |  |  |  |  |  |  |  |  |  |  |  |  |  |  |  |
| --- | --- | --- | --- | --- | --- | --- | --- | --- | --- | --- | --- | --- | --- | --- | --- | --- |
| STUDY | Ground truth categories |  |  |  |  |  |  |  |  |  | GRAND TOTAL BY STUDY<br>(n=17013, n=9462) |  |  |  |  |  |
|  | no. (%) |  |  |  |  |  |  |  |  |  |  |  |  |  |  |  |
|  | Normal (n=11630, n=6092) |  |  | Gray Zone (n=3586, n=2314) |  |  | Precancer+ (n=1797, n=1056) |  |  |  | no. (%) |  |  |  |  |  |
|  | # images |  | # women | # images |  | # women | # images |  | # women | # images |  | # women | # images |  | # women |  |
| Train Set |  |  |  |  |  |  |  |  |  |  |  |  |  |  |  |  |
| NHS | 1887 | (77.6%) | 946 | (74.4%) | 330 | (15.3%) | 165 | (11.9%) | 206 | (19.0%) | 104 | (16.4%) | 2423 | (42.7%) | 1215 | (36.8%) |
| ALTS | 387 | (15.9%) | 194 | (15.3%) | 853 | (39.6%) | 430 | (30.9%) | 434 | (40.1%) | 218 | (34.3%) | 1674 | (29.5%) | 842 | (25.5%) |
| CVT | 88 | (3.6%) | 88 | (6.9%) | 336 | (15.6%) | 335 | (24.1%) | 121 | (11.2%) | 119 | (18.7%) | 545 | (9.6%) | 542 | (16.4%) |
| Biop | 35 | (1.4%) | 13 | (1.0%) | 192 | (8.9%) | 88 | (6.3%) | 164 | (15.2%) | 79 | (12.4%) | 391 | (6.9%) | 180 | (5.5%) |
| D Biop | 35 | (1.4%) | 31 | (2.4%) | 444 | (20.6%) | 374 | (26.9%) | 157 | (14.5%) | 116 | (18.2%) | 636 | (11.2%) | 521 | (15.8%) |
| TOTAL | 2432 | (100.0%) | 1272 | (100.0%) | 2155 | (100.0%) | 1392 | (100.0%) | 1082 | (100.0%) | 636 | (100.0%) | 5669 | (100.0%) | 3300 | (100.0%) |
| (a) | 42.9% |  | 38.5% |  | 38.0% |  | 42.2% |  | 19.1% |  | 19.3% |  | 100.0% |  | 100.0% |  |
| (b) |  |  |  |  |  |  |  |  |  |  |  |  | 33.3% |  | 34.9% |  |
| Validation Set |  |  |  |  |  |  |  |  |  |  |  |  |  |  |  |  |
| NHS | 291 | (76.0%) | 146 | (71.6%) | 55 | (15.1%) | 28 | (12.3%) | 34 | (19.2%) | 17 | (16.7%) | 380 | (41.1%) | 191 | (35.8%) |
| ALTS | 65 | (17.0%) | 33 | (16.2%) | 142 | (39.0%) | 71 | (31.1%) | 72 | (40.7%) | 36 | (35.3%) | 279 | (30.2%) | 140 | (26.2%) |
| CVT | 19 | (5.0%) | 19 | (9.3%) | 53 | (14.6%) | 53 | (23.2%) | 17 | (9.6%) | 17 | (16.7%) | 89 | (9.6%) | 89 | (16.7%) |
| Biop | 4 | (1.0%) | 2 | (1.0%) | 35 | (9.6%) | 14 | (6.1%) | 29 | (16.4%) | 13 | (12.7%) | 68 | (7.4%) | 29 | (5.4%) |
| D Biop | 4 | (1.0%) | 4 | (2.0%) | 79 | (21.7%) | 62 | (27.2%) | 25 | (14.1%) | 19 | (18.6%) | 108 | (11.7%) | 85 | (15.9%) |
| TOTAL | 383 | (100.0%) | 204 | (100.0%) | 364 | (100.0%) | 228 | (100.0%) | 177 | (100.0%) | 102 | (100.0%) | 924 | (100.0%) | 534 | (100.0%) |
| (a) | 41.5% |  | 38.2% |  | 39.4% |  | 42.7% |  | 19.2% |  | 19.1% |  | 100.0% |  | 100.0% |  |
| (b) |  |  |  |  |  |  |  |  |  |  |  |  | 5.4% |  | 5.6% |  |
| Test Set 1 |  |  |  |  |  |  |  |  |  |  |  |  |  |  |  |  |
| NHS | 5930 | (77.4%) | 2974 | (74.1%) | 108 | (15.3%) | 55 | (11.9%) | 70 | (19.1%) | 35 | (16.2%) | 6108 | (69.9%) | 3064 | (65.3%) |
| ALTS | 1240 | (16.2%) | 622 | (15.5%) | 285 | (40.3%) | 143 | (31.0%) | 146 | (39.8%) | 73 | (33.8%) | 1671 | (19.1%) | 838 | (17.9%) |
| CVT | 280 | (3.7%) | 280 | (7.0%) | 110 | (15.6%) | 110 | (23.8%) | 42 | (11.4%) | 42 | (19.4%) | 432 | (4.9%) | 432 | (9.2%) |
| Biop | 94 | (1.2%) | 44 | (1.1%) | 60 | (8.5%) | 29 | (6.3%) | 55 | (15.0%) | 27 | (12.5%) | 209 | (2.4%) | 100 | (2.1%) |
| D Biop | 116 | (1.5%) | 92 | (2.3%) | 144 | (20.4%) | 125 | (27.1%) | 54 | (14.7%) | 39 | (18.1%) | 314 | (3.6%) | 256 | (5.5%) |
| TOTAL | 7660 | (100.0%) | 4012 | (100.0%) | 707 | (100.0%) | 462 | (100.0%) | 367 | (100.0%) | 216 | (100.0%) | 8734 | (100.0%) | 4690 | (100.0%) |
| (a) | 87.7% |  | 85.5% |  | 8.1% |  | 9.9% |  | 4.2% |  | 4.6% |  | 100.0% |  | 100.0% |  |
| (b) |  |  |  |  |  |  |  |  |  |  |  |  | 51.3% |  | 49.6% |  |
| Test Set 2 |  |  |  |  |  |  |  |  |  |  |  |  |  |  |  |  |
| NHS | 902 | (78.1%) | 452 | (74.8%) | 54 | (15.0%) | 27 | (11.6%) | 34 | (19.9%) | 17 | (16.7%) | 990 | (58.7%) | 496 | (52.9%) |
| ALTS | 187 | (16.2%) | 94 | (15.6%) | 144 | (40.0%) | 72 | (31.0%) | 72 | (42.1%) | 36 | (35.3%) | 403 | (23.9%) | 202 | (21.5%) |
| CVT | 37 | (3.2%) | 37 | (6.1%) | 56 | (15.6%) | 56 | (24.1%) | 17 | (9.9%) | 17 | (16.7%) | 110 | (6.5%) | 110 | (11.7%) |
| Biop | 14 | (1.2%) | 7 | (1.2%) | 28 | (7.8%) | 15 | (6.5%) | 27 | (15.8%) | 13 | (12.7%) | 69 | (4.1%) | 35 | (3.7%) |
| D Biop | 15 | (1.3%) | 14 | (2.3%) | 78 | (21.7%) | 62 | (26.7%) | 21 | (12.3%) | 19 | (18.6%) | 114 | (6.8%) | 95 | (10.1%) |
| TOTAL | 1155 | (100.0%) | 604 | (100.0%) | 360 | (100.0%) | 232 | (100.0%) | 171 | (100.0%) | 102 | (100.0%) | 1686 | (100.0%) | 938 | (100.0%) |
| (a) | 68.5% |  | 64.4% |  | 21.4% |  | 24.7% |  | 10.1% |  | 10.9% |  | 100.0% |  | 100.0% |  |
| (b) |  |  |  |  |  |  |  |  |  |  |  |  | 9.9% |  | 9.9% |  |
| GRAND TOTAL BY GROUND TRUTH |  |  |  |  |  |  |  |  |  |  |  |  |  |  |  |  |
| no. (%) | 11630 |  | 6092 |  | 3586 |  | 2314 |  | 1797 |  | 1056 |  | 17013 |  | 9462 |  |
|  | (68.4%) |  | (64.4%) |  | (21.1%) |  | (24.5%) |  | (10.6%) |  | (11.2%) |  | (100.0%) |  | (100.0%) |  |

